# Referral of febrile children in resource-constrained community settings in Asia (Spot Sepsis) – a multi-country, prospective, cohort study

**DOI:** 10.1101/2025.10.12.25337822

**Authors:** Arjun Chandna, Constantinos Koshiaris, Raman Mahajan, Riris Adono Ahmad, Dinh Thi Van Anh, Khalid Shams Choudhury, Suy Keang, Phung Nguyen The Nguyen, Sayaphet Rattanavong, Souphaphone Vannachone, Spot Sepsis Investigator Group, Chris Painter, Mikhael Yosia, Naomi Waithira, Mohammad Yazid Abdad, Janjira Thaipadungpanit, Paul Turner, Phan Huu Phuc, Dinesh Mondal, Mayfong Mayxay, Bui Thanh Liem, Elizabeth A Ashley, Eggi Arguni, Rafael Perera-Salazar, Melissa Richard-Greenblatt, Yoel Lubell, Sakib Burza

## Abstract

In resource-constrained community settings, distinguishing which febrile children require referral is a major unmet need. Current WHO danger signs lack accuracy, resulting in missed severe illness and unnecessary referrals. We developed and validated simple clinical prediction models using data from 3,405 children aged 1-59 months presenting with community-acquired acute febrile illnesses to seven hospitals across Bangladesh, Cambodia, Indonesia, Laos, and Viet Nam. Cambodian data were held-out for external validation. All models outperformed WHO criteria to predict progression to severe febrile illness (death or organ support) within two days (sensitivity=0.56, 95%CI=0.42-0.69; specificity=0.83, 95%CI=0.78-0.87). Incorporating pulse oximetry or the host biomarker sTREM1 further enhanced sensitivity (0.89, 95%CI=0.79-0.97) vs. clinical features alone (0.75, 95%CI=0.62-0.86). The pulse oximetry-based model achieved these gains while improving specificity, concomitantly reducing referral rates three-fold. These approaches appear cost-effective and could transform referral practices for febrile children in resource-constrained community settings. They warrant evaluation in randomised controlled trials.

## INTRODUCTION

Infectious diseases account for the majority of the 2.5 million deaths that occur each year among children aged 1-59 months.^1^ Many deaths happen in community settings, either because a child does not attend health services, presents too late, referral is impractical, or there is a failure to recognise impending critical illness.^2–4^ Early identification of children at risk of life-threatening infection is difficult.^5,6^ Most Early Warning Scores (EWS) rely on combinations of abnormal vital signs (for example, fever, tachycardia, and tachypnoea) but these are dynamic, prone to confounding, and poorly sensitive and specific for identifying early stages of sepsis.^7,8^

In resource-constrained community settings, current World Health Organization (WHO) Integrated Management of Childhood Illnesses (IMCI) guidelines do not advocate the use of vital signs to assess illness severity.^9,10^ Instead, hospital referral is prompted by clinical danger signs, such as convulsions, intractable vomiting, lethargy, or prostration. Accuracy of these indicators is sub-optimal and they are prone to considerable inter-observer variability.^7,11–13^ Neither vital nor danger signs reliably stratify risk in common childhood infections, underscoring the need for better prognostic tools.^14^ Such tools would be particularly valuable in remote and conflict-affected settings, where capacity for safety-netting is typically minimal while the opportunity cost of unnecessary referral is high.

One option, spotlighted by the Covid-19 pandemic, is expansion of pulse oximetry to primary care settings. Championed as the ‘fifth vital sign’,^15^ hypoxaemia predicts poor outcome in childhood illnesses including pneumonia, meningitis, malaria, and malnutrition, and identifies large numbers of children requiring hospital referral missed by IMCI guidelines alone.^16–20^ Despite this promise, pulse oximetry remains under-utilised in community settings. Barriers include availability, cost, and usability of appropriately sized probes, accuracy across skin tones, and staff capacity and workload: oximetry takes time, especially in young children where measurement can be challenged by movement or crying.^21–23^ Implementation by community health workers is likely to be especially challenging.

Another strategy is the integration of clinical assessment with host biomarker testing.^5,24^ Circulating markers of immune and endothelial activation predict disease severity agnostic to pathogen aetiology and, in certain contexts, have outperformed pulse oximetry.^25–29^ We recently reported promising prognostic performance of host biomarkers at the community level,^30^ and an operational evaluation among village malaria workers in Cambodia further demonstrated the feasibility of this approach.^31^ If effective, such strategies could capitalise on point-of-care testing capacity developed among community healthcare worker networks over the preceding decades by control programmes for diseases such as malaria and HIV.^32^

In this study, our objective was to develop and validate clinical prediction models to guide referral of febrile children from resource-constrained community contexts and decentralised models of care. Specifically, we aimed to develop models comprising simple clinical parameters (for example, vital and danger signs) to predict disease progression and explore the added utility of including pulse oximetry and host biomarker tests.

## METHODS

### Study design and setting

Spot Sepsis was a multi-country, prospective, cohort study conducted in seven hospitals across Bangladesh, Cambodia, Indonesia, Lao PDR (Laos), and Viet Nam. Study sites were purposefully chosen to represent facilities serving rural populations and providing a first point of access to the formal healthcare sector, as a proxy for the ultimate intended use-case in community care settings (appendix p2-3). Data from six sites were used to derive the clinical prediction models, with the site in Cambodia prespecified for held-out external geographic validation. This split was pragmatic, based on the ability to conduct recruitment for longer in Cambodia.

### Study participants

Children (aged > 28 days and < 60 months) presenting with a febrile illness (axillary temperature ≥ 37.5°C or < 35.5°C or history of fever in the preceding 24 hours)^9,33^ of ≤ 14 days were eligible for inclusion. Exclusion criteria were prior admission to any health facility during the current illness, receipt of > 15 minutes of parenteral treatment (intravenous, intramuscular, or nebulised medication, intravenous fluids, or supplemental oxygen) prior to screening,^34^ presentation within three days of routine immunisations, trauma as the reason for attendance, or specific known comorbidities (chronic infection [e.g. viral hepatitis or tuberculosis], immunosuppression [e.g. HIV or oncological conditions], or active [symptomatic or currently medicated] cardiorespiratory conditions). Participants could only be enrolled once.

### Screening and enrolment

Patients were screened during daytime working hours. Screening was stratified by admission status. Admissions could occur via the emergency or outpatient departments. Inpatients were screened consecutively upon arrival at the emergency department or inpatient ward. Children admitted from the outpatient department typically did not receive treatment before arrival on the inpatient ward. Children attending the emergency department typically received treatment promptly and were either admitted or kept for a period of observation. Thus, all patients presenting to the emergency department (both those admitted and those kept for a period of observation) were considered inpatients.

Due to high outpatient numbers, outpatient screening was randomised. Outpatients to be approached were randomly allocated by computer-generated random number tables with the preceding week’s routinely collected hospital attendance data providing the sampling frame. Screening of outpatients occurred prior to assessment by the clinical team, with three outpatients targeted for recruitment each week at each site. On rare occasions when the health worker decided to admit a child just recruited into the outpatient strata, the participant was transferred to the inpatient strata and the next patient specified on the random number table was approached for screening.

### Data collection

Trained study personnel measured vital signs (including SpO_2_ via pulse oximetry) and anthropometrics, assessed clinical signs (including WHO danger signs), and collected venous blood samples and nasopharyngeal swabs at enrolment (Table S1; appendix p4).

Demographic information and perinatal, past medical, and illness histories were collected via interview with the participant’s caregiver. All data were entered onto electronic case record forms using Android tablets via Open Data Kit Collect software. Participants were provided with routine care by their treating clinician. When feasible, the study supported collection and processing of peripheral blood cultures at the discretion of the clinical team.

Participants were followed-up on days 2 and 28 after enrolment, with additional follow-up on day 1 and at discharge for inpatients. Follow-up was conducted in-person if the participant remained admitted at the study site or by telephone if they had been discharged. Study monitoring was conducted by the Clinical Trials Support Group at the Mahidol Oxford Tropical Medicine Research Unit (MORU) in Bangkok, Thailand.

### Clinical parameters and host biomarkers

Clinical parameters useful for the prognostication of paediatric febrile illness were identified through systematic review of the literature.^35^ Feasibility of measurement in resource-constrained community settings was considered,^36^ with only the most practicable parameters retained. Prioritisation and standardisation followed guidance set out by the Pediatric Sepsis Predictors Standardization working group,^37^ operationalised through Standard Operating Procedures (SOPs) on which all study personnel were trained prior to beginning data collection. Adherence to SOPs was assessed at Site Initiation Visits and regular monitoring visits. Study equipment was procured centrally and distributed to sites.

Host response biomarkers were shortlisted following review of the literature and consultation with domain experts (Table S2; appendix p5). In primary care, where the cause of infection is typically unknown at the time of presentation, biomarkers that are predictive across a range of pathogens are essential for risk stratification. Therefore, biomarkers implicated in final common pathways to severe febrile illness and sepsis were prioritised.^38–41^ Endothelial activation markers included ANG-1, ANG-2, and soluble FLT-1 (sFLT-1; also known as sVEGFR-1). Immune activation markers included CHI3L1, CRP, IP-10 (also known as CXCL10), IL-1ra, IL-6, IL-8, IL-10, PCT, soluble TNF-R1 (sTNF-R1), soluble TREM1 (sTREM1), and soluble uPAR (suPAR). Lactate, glucose, and haemoglobin were also included because they are easily measured using inexpensive rapid tests, well-known to clinicians, have prognostic value,^35^ and recommended in paediatric sepsis guidelines.^33,42,43^

### Laboratory procedures

Venous blood samples and nasopharyngeal swabs were processed immediately. Complete blood counts were conducted on-site, and peripheral blood cultures were processed at in-country laboratories. Aliquots of whole blood, EDTA-plasma, fluoride-oxalate-plasma, and universal transport medium (UTM) were stored at −20°C or lower. These samples were then transported at −80°C to the MORU laboratories in Bangkok, Thailand, for further analysis and biobanking.

Biomarker concentrations in EDTA-plasma were measured using the Simple Plex Ella microfluidic platform (ProteinSimple, San Jose, CA, USA) and the suPARnostic ELISA (ViroGates, Copenhagen, Denmark), as outlined in the appendix (Table S3; p6). Glucose (GLUC3, Roche Diagnostics, Mannheim, Germany) and lactate (LACT2, Roche Diagnostics, Germany) concentrations were quantified in fluoride-oxalate-plasma.

Nucleic acid was extracted from whole blood using the MagNA Pure 24 instrument and Total NA Isolation Kit (Roche Diagnostics, Indianapolis, IN, USA), following the manufacturer’s instructions. Real-time polymerase chain reaction (RT-PCR) multiplex assays were used to detect viral (chikungunya, dengue, Japanese encephalitis, and Zika) and bacterial (*Leptospira* spp., *Orientia tsutsugamushi*, and *Rickettsia* spp.) targets in whole blood. Respiratory pathogen targets were detected directly from nasopharyngeal swabs using the FilmArray RP2 panel (BioFire Diagnostics, Salt Lake City, UT, USA), except for samples from Cambodia, which were processed for influenza A/B and respiratory syncytial virus (RSV) using the FTD FLU/HRSV assay (Siemens, Erlangen, Germany). Specimens from all sites were tested using an in-house multiplex RT-PCR assay to detect SARS-CoV-2 in nasopharyngeal swabs, based on the E and N genes, as previously described.^44^

### Outcome measures

The primary outcome was development of severe febrile illness within two days of enrolment, defined as death or receipt of organ support (mechanical ventilation, non-invasive ventilation, inotropic therapy, or renal replacement therapy).

Recognising that illness severity is a continuum, participant outcomes were also classified on a categorical scale (Table S4; appendix p7). This was used to further assess the performance of the prediction models and provide insight into the relative importance of misclassifications.

### Sample size

The Covid-19 pandemic, declared in the same week that recruitment began at the first site, delayed initiation of other sites and slowed enrolment. Study duration (and thus sample size) was determined by available resources. Prior to study commencement, the methods of Riley et al. were followed and using an anticipated outcome prevalence of 1%, conservative R^2^ Nagelkerke of 0.15, and shrinkage factor of 0.9, it was estimated that six events per parameter (EPP) would be required to derive the prediction models.^45^ Accordingly, the maximum number of candidate predictors that could be used to derive the models was 16, whilst minimising the risk of overfitting.

### Statistical methods

Categorical and continuous variables were summarised using descriptive statistics for both the derivation and validation cohorts, and compared between participants according to their outcome status, using the Wilcoxon rank sum test, Pearson’s X^2^ test, or Fisher’s exact test as appropriate. Site-specific outpatient weights were applied to adjust for unequal probabilities of selection in the sample, arising due to random sampling of outpatients (Table S5; appendix p8-9), to ensure the study population was representative of all eligible children presenting to the hospital and that the outcome prevalence reflected that which might be observed in community care settings.^46,47^

Candidate predictors were prespecified using domain knowledge and literature review (Table S6; appendix p10). The relationship between each continuous candidate predictor and the primary outcome was explored by visually inspecting a smoothing curve to identify non-linear patterns and by examining the overall calibration of the model. Transformations were considered only for serious violations of linearity which were substantially affecting model calibration. There were few missing data (≤ 6.0%) for all predictors except haemoglobin, where 16.9% (574/3,405) of participants had missing data, as complete blood counts were collected at the discretion of the treating clinical team. For all predictors, missing observations were replaced with their median value, conditional on outcome status.

To build the *clinical model*, all 11 prespecified candidate clinical predictors (excluding SpO_2_) were entered into the model and backward stepwise weighted logistic regression used to identify the most parsimonious model. This process was repeated to build the *pulse oximetry model*, starting with all prespecified candidate clinical predictors, this time including SpO_2_. To identify the most promising biomarker, *clinical-biomarker models* were developed using backward stepwise weighted logistic regression with all prespecified candidate clinical predictors (with and without SpO_2_) and each of the 17 biomarkers in turn. The models were internally validated using bootstrapping, with 800 samples being drawn with replacement, to adjust for optimism. A sensitivity analysis excluding the northern Viet Nam site (n=612), which departed from the ideal rural target site profile and where outpatient weighting was derived using different methodology (Table S5; appendix p8-9), was performed. The *clinical-biomarker model* with highest discrimination was carried forward for external validation. Overall, a total of four models (*clinical model*, *pulse oximetry model*, a *clinical-biomarker model* and a *combined model* including the biomarker and SpO_2_) were taken forward for validation in the validation cohort.

Models were applied to the validation dataset and the predicted probabilities estimated. Model performance was evaluated in terms of discrimination and calibration. Discrimination – the ability of a model to distinguish between individuals who did and did not experience the outcome – was assessed using the weighted AUC, with values ranging from 0.5 (no discrimination) to 1.0 (perfect discrimination). Calibration – the agreement between predicted and observed risks – was assessed using the weighted calibration intercept and slope. An ideal model has an intercept of zero (with positive values indicating overall risk underestimation and negative values indicating overestimation) and a slope of one (where slopes > 1 suggest underestimation in low-risk individuals and overestimation in high-risk individuals, and slopes < 1 indicate the opposite pattern).

Clinical utility of each model was assessed by evaluating classification (sensitivity, specificity, negative likelihood ratio, and positive likelihood ratio) at clinically relevant cut-points (predicted probabilities of severe febrile illness). For each model, the distribution of predicted probabilities and triage groups was examined in relation to the categorical outcome scale, to better understand the potential implications of misclassifications. Finally, the characteristics of participants who developed severe febrile illness but were missed by the models were explored, in order to understand model vulnerabilities and determine what adjustments may lead to improved model performance.

### Cost-effectiveness analyses

A cost-effectiveness analysis of the different models for the triage of febrile children was conducted, set in the context of Bangladesh, which was the study site most closely aligned with the intended use setting for the referral tool (Figure S1; appendix p11). The analyses were parameterised using primary data from the study and published estimates available in the literature (Table S7; appendix p12). Two CETs were explored: $2,551 per disability adjusted life year (DALY) averted (approximating Bangladesh’s 2023 GDP per capita)^48,49^ and a more conservative CET of $459/DALY averted obtained from Woods and colleagues (inflated to 2024 values).^50^ A sensitivity analysis was conducted to determine the maximum referral cost at which the different models might remain cost-effective, for both of the analysed CETs.

### Ethics, funding, inclusion, and reporting

Caregivers of all participants provided informed written consent. The study was prospectively registered on ClinicalTrials.gov (NCT04285021) and received ethical approval from the sponsors and ethical review boards in all participating countries (Table S8; appendix p13). The study was co-funded by Médecins Sans Frontières, Spain (MSF) and Wellcome. MSF maintained a sponsor-investigator role for the study. Wellcome had no role in study design, data collection, data analysis, data interpretation, writing of the report, or decision to submit for publication.

The study was designed and implemented collaboratively by investigators from all participating countries, based on feedback from stakeholders, healthcare providers, and the Young Persons Advisory Group at Angkor Hospital for Children, Cambodia. Progress was reviewed at monthly investigator meetings, where feedback from the site teams was discussed.

The study is reported in accordance with the Transparent Reporting of a multivariable prediction model for Individual Prognosis Or Diagnosis (TRIPOD) guidelines (Table S9; appendix p14).^51^

## RESULTS

### Study population

A total of 11,962 children were screened between 5 March 2020 and 4 November 2022. Of these, 3,998 were eligible (3,998/11,962; 33.4%) and 3,423 were recruited (575/3,998; 14.4% refusal rate). There were two withdrawals and 16 children with incomplete follow-up data, leaving 3,405 participants for further analyses (Figure S2; appendix p15).

Median age was 16.8 months (interquartile range [IQR] 8.7-31.0) and 59.6% (2,029/3,405) of the cohort were male. Malnutrition was prevalent: 17.2% (585/3,393) of children were wasted (weight-for-height z-score <-2) and 19.5% (664/3,401) were stunted (height-for-age z-score <-2). Of the 585 children with Global Acute Malnutrition (GAM), 248 (42.4%) had Severe Acute Malnutrition (SAM; weight-for-height z-score <-3). Median symptom duration prior to presentation was three days (IQR 2-4). Acute respiratory infections were the most common reason for presentation, followed by diarrhoeal syndromes and undifferentiated febrile illnesses. A quarter of participants had a microbiological cause for their infection identified (898/3,405; 26.4%; Table S10; appendix p16). Most children lived close to the hospital (2,777/3,405; 81.6% within an hour). In total, 1,342 participants (1,342/3,405; 39.4%) had received care in the community at an earlier point in their illness: of these, 193 (193/3,405; 5.7%) had received parenteral treatment and none had been admitted.

Baseline characteristics were largely balanced across derivation and validation cohorts (Table 1), although the validation cohort were younger (median age: 13.1 months vs. 18.7 months) and a higher proportion had received parenteral treatment in the community at an earlier point in their illness (109/824; 13.2% vs. 84/2,581; 3.3%). Baseline biomarker concentrations were similar across the cohorts. There were a greater proportion of confirmed viral infections in the derivation cohort (709/2,581; 27.4% vs. 155/824; 18.7%). Bacteraemia rates were similar (12/781; 1.5% vs. 7/411; 1.7%), although blood cultures were collected more frequently from inpatients in the validation cohort (411/644; 63.8% vs. 781/2,080; 37.5%).

**TABLE 1.** Baseline characteristics of the derivation and validation cohort, stratified by whether a child progressed to develop severe febrile illness.

| Characteristic | Derivation cohort |  |  |  | Validation cohort |  |  |  |
| --- | --- | --- | --- | --- | --- | --- | --- | --- |
|  | Overall<br>N = 2,581<sup>1</sup> | Non-severe<br>N = 2,484<sup>1</sup> | Severe<br>N = 97<sup>1</sup> | p-value<sup>2</sup> | Overall<br>N = 824<sup>1</sup> | Non-severe<br>N = 788<sup>1</sup> | Severe<br>N = 36<sup>1</sup> | p-value<sup>2</sup> |
| **Demographics and background** |  |  |  |  |  |  |  |  |
| Age (months) | 18.7 (9.3, 32.9) | 18.9 (9.5, 33.1) | 6.9 (3.2, 25.8) | < 0.0001 | 13.1 (7.5, 24.4) | 13.5 (8.1, 24.9) | 3.4 (1.8, 6.2) | < 0.0001 |
| Male sex | 1,577 (61%) | 1513 (61%) | 64 (66%) | 0.32 | 452 (55%) | 428 (54%) | 24 (67%) | 0.15 |
| Known comorbidity | 65 (2.5%) | 63 (2.5%) | 2 (2.1%) | 1.00 | 37 (4.5%) | 37 (4.7%) | 0 (0%) | 0.40 |
| Recent hospitalisation<sup>a,*</sup> | 277 (11%) | 265 (11%) | 12 (13%) | 0.58 | 152 (19%) | 147 (19%) | 5 (14%) | 0.46 |
| **Anthropometrics** |  |  |  |  |  |  |  |  |
| Wasted (WHZ < -2)* | 474 (18%) | 445 (18%) | 29 (30%) | 0.0030 | 111 (14%) | 106 (14%) | 5 (14%) | 0.80 |
| Stunted (HAZ < -2)* | 484 (19%) | 462 (19%) | 22 (23%) | 0.31 | 180 (22%) | 167 (21%) | 13 (36%) | 0.035 |
| MUAC-for-age z-score<sup>b,*</sup> | -0.2 (-1.1, 0.7) | -0.2 (-1.1, 0.7) | -0.5 (-1.4, 0.7) | 0.12 | -0.5 (-1.2, 0.2) | -0.5 (-1.1, 0.2) | -0.8 (-1.7, 0.3) | 0.40 |
| **Illness history** |  |  |  |  |  |  |  |  |
| Duration of illness (days) | 3.0 (2.0, 4.0) | 3.0 (2.0, 4.0) | 3.0 (2.0, 5.0) | 0.020 | 3.0 (2.0, 4.0) | 3.0 (2.0, 4.0) | 3.5 (3.0, 5.0) | 0.051 |
| Care prior to presentation<sup>c</sup> | 864 (33%) | 829 (33%) | 35 (36%) | 0.58 | 478 (58%) | 457 (58%) | 21 (58%) | 0.97 |
| Travel time ≤ 1 hour<sup>d</sup> | 2,227 (86%) | 2,150 (87%) | 77 (79%) | 0.044 | 550 (67%) | 532 (68%) | 18 (50%) | 0.029 |
| **Presenting syndrome** |  |  |  |  |  |  |  |  |
| URTI | 795 (31%) | 773 (31%) | 22 (23%) | 0.077 | 326 (40%) | 309 (39%) | 17 (47%) | 0.34 |
| LRTI | 930 (36%) | 877 (35%) | 53 (55%) | 0.0001 | 417 (51%) | 384 (49%) | 33 (92%) | < 0.0001 |
| Diarrhoeal | 397 (15%) | 386 (16%) | 11 (11%) | 0.26 | 249 (30%) | 245 (31%) | 4 (11%) | 0.011 |
| No focus | 372 (14%) | 366 (15%) | 6 (6.2%) | 0.019 | 155 (19%) | 153 (19%) | 2 (5.6%) | 0.037 |
| Neurological | 372 (14%) | 359 (14%) | 13 (13%) | 0.77 | 58 (7.0%) | 57 (7.2%) | 1 (2.8%) | 0.51 |
| **WHO danger signs** |  |  |  |  |  |  |  |  |
| Any danger sign present* | 1,246 (48%) | 1,171 (47%) | 75 (78%) | < 0.0001 | 361 (44%) | 341 (43%) | 20 (56%) | 0.15 |
| Prostration | 199 (7.7%) | 155 (6.2%) | 44 (45%) | < 0.0001 | 41 (5.0%) | 37 (4.7%) | 4 (11%) | 0.098 |
| Intractable vomiting* | 604 (23%) | 564 (23%) | 40 (41%) | < 0.0001 | 83 (10%) | 81 (10%) | 2 (5.6%) | 0.57 |
| Convulsions* | 373 (14%) | 360 (15%) | 13 (14%) | 0.79 | 60 (7.3%) | 59 (7.5%) | 1 (2.8%) | 0.51 |
| Lethargy* | 520 (20%) | 478 (19%) | 42 (44%) | < 0.0001 | 305 (37%) | 286 (36%) | 19 (53%) | 0.045 |
| **Vital signs** |  |  |  |  |  |  |  |  |
| Heart rate (bpm)* |  |  |  |  |  |  |  |  |
| 1 to 12 months | 154.0 (140.0, 171.0) | 153.0 (140.0, 170.0) | 170.0 (152.0, 184.0) | < 0.0001 | 159.0 (143.0, 176.0) | 157.0 (142.0, 173.0) | 177.0 (161.0, 186.0) | < 0.0001 |
| 12 to 60 months | 140.0 (126.0, 157.0) | 140.0 (126.0, 156.0) | 159.0 (140.0, 179.5) | < 0.0001 | 140.0 (128.0, 162.0) | 140.0 (128.0, 162.0) | 167.0 (146.0, 172.0) | 0.17 |
| Respiratory rate (bpm)* |  |  |  |  |  |  |  |  |
|  | Overall<br>N = 2,581<sup>1</sup> | Non-severe<br>N = 2,484<sup>1</sup> | Severe<br>N = 97<sup>1</sup> | p-value<sup>2</sup> | Overall<br>N = 824<sup>1</sup> | Non-severe<br>N = 788<sup>1</sup> | Severe<br>N = 36<sup>1</sup> | p-value<sup>2</sup> |
| *1 to 12 months* | 45.0 (38.0, 55.0) | 44.0 (38.0, 53.0) | 60.0 (50.0, 68.0) | < 0.0001 | 44.0 (37.0, 54.0) | 44.0 (36.0, 52.0) | 58.0 (46.0, 64.0) | < 0.0001 |
| *12 to 60 months* | 35.0 (30.0, 41.0) | 35.0 (30.0, 41.0) | 39.0 (31.5, 55.0) | 0.016 | 38.0 (32.0, 45.0) | 38.0 (32.0, 45.0) | 54.0 (50.0, 67.0) | 0.032 |
| Oxygen saturation (%)* | 98.0 (97.0, 99.0) | 98.0 (97.0, 99.0) | 97.0 (95.0, 98.0) | 0.0004 | 99.0 (98.0, 100.0) | 99.0 (98.0, 100.0) | 96.0 (94.5, 97.5) | < 0.0001 |
| Axillary temperature (°C)* | 37.7 (37.0, 38.4) | 37.7 (37.0, 38.4) | 38.0 (37.2, 38.6) | 0.098 | 37.3 (36.7, 38.1) | 37.3 (36.7, 38.1) | 37.0 (36.5, 37.5) | 0.019 |
| CRT > 2 seconds | 180 (7.0%) | 153 (6.2%) | 27 (28%) | < 0.0001 | 14 (1.7%) | 12 (1.5%) | 2 (5.6%) | 0.12 |
| Not alert<sup>e</sup> | 58 (2.2%) | 41 (1.7%) | 17 (18%) | < 0.0001 | 28 (3.4%) | 26 (3.3%) | 2 (5.6%) | 0.35 |
| **Endothelial activation markers** |  |  |  |  |  |  |  |  |
| ANG-1 (pg/ml)* | 6,396.5<br>(3,387.5, 11,645.0) | 6,411.0<br>(3,387.0, 11,607.0) | 6,049.0<br>(3,426.0, 13,382.0) | 1.00 | 5,686.5<br>(3,454.0, 9,139.0) | 5,713.5<br>(3,454.0, 9,240.0) | 4,621.0<br>(3,440.5, 7,421.5) | 0.24 |
| ANG-2 (pg/ml)* | 1,330.0<br>(998.5, 1,844.0) | 1,320.0<br>(993.0, 1,802.0) | 2,343.0<br>(1,317.0, 4,608.0) | < 0.0001 | 1,455.5<br>(1,090.0, 2,051.0) | 1,427.0<br>(1,081.0, 1,965.0) | 3,008.0<br>(2,119.0, 4,254.5) | < 0.0001 |
| sFLT-1 (pg/ml)* | 183.0<br>(149.0, 231.0) | 181.0<br>(148.0, 227.0) | 258.0<br>(196.0, 369.0) | < 0.0001 | 196.0<br>(156.0, 245.0) | 194.5<br>(154.0, 242.0) | 260.5<br>(227.0, 415.5) | < 0.0001 |
| **Immune activation markers** |  |  |  |  |  |  |  |  |
| CHI3L1 (ng/ml)* | 23.8<br>(15.7, 37.7) | 23.5<br>(15.5, 36.9) | 37.6<br>(22.8, 62.6) | < 0.0001 | 28.9<br>(20.2, 43.1) | 28.5<br>(20.1, 42.5) | 38.2<br>(26.9, 72.8) | 0.0037 |
| CRP (mg/l)* | 13.2<br>(3.2, 49.3) | 12.8<br>(3.2, 47.8) | 31.4<br>(6.8, 150.5) | 0.0001 | 17.3<br>(4.8, 55.3) | 17.5<br>(4.8, 55.3) | 11.8<br>(2.1, 55.5) | 0.31 |
| IL-1ra (pg/ml)* | 2,036.0<br>(980.0, 4,875.0) | 1,983.0<br>(967.0, 4,764.0) | 4,117.0<br>(1,756.0, 14,784.0) | < 0.0001 | 1,755.0<br>(789.0, 4,429.0) | 1,781.0<br>(806.0, 4,440.0) | 1,505.0<br>(728.5, 3,509.5) | 0.32 |
| IL-6 (pg/ml)* | 18.4<br>(7.7, 46.3) | 17.6<br>(7.6, 43.5) | 73.7<br>(20.9, 320.0) | < 0.0001 | 16.9<br>(7.2, 46.9) | 16.8<br>(7.2, 46.9) | 17.8<br>(7.7, 48.7) | 0.80 |
| IL-8 (pg/ml)* | 15.4<br>(9.2, 27.5) | 15.1<br>(9.1, 26.8) | 35.7<br>(13.1, 129.0) | < 0.0001 | 15.6<br>(9.3, 27.6) | 15.2<br>(9.1, 26.4) | 23.7<br>(16.4, 38.8) | 0.0035 |
| IL-10 (pg/ml)* | 19.3<br>(10.5, 43.6) | 19.1<br>(10.3, 42.6) | 34.7<br>(15.5, 83.3) | < 0.0001 | 20.5<br>(10.8, 43.6) | 20.1<br>(10.8, 43.6) | 23.8<br>(12.0, 43.4) | 0.62 |
| IP-10 (pg/ml)* | 852.5<br>(387.5, 1,882.0) | 857.0<br>(394.0, 1,895.0) | 745.0<br>(248.0, 1,381.0) | 0.023 | 727.0<br>(368.0, 1,858.0) | 754.5<br>(383.0, 1,929.0) | 371.0<br>(218.0, 692.0) | 0.0001 |
| PCT (ng/ml)* | 0.3<br>(0.2, 0.7) | 0.3<br>(0.2, 0.7) | 1.0<br>(0.4, 6.1) | < 0.0001 | 0.4<br>(0.2, 1.1) | 0.4<br>(0.2, 1.1) | 0.7<br>(0.4, 1.6) | 0.0072 |
| sTNF-R1 (pg/ml)* | 1,559.0<br>(1,244.0, 2,022.0) | 1,546.0<br>(1,240.0, 1,992.0) | 2,166.0<br>(1,549.0, 3,542.0) | < 0.0001 | 1,643.5<br>(1,306.0, 2,153.0) | 1,627.0<br>(1,297.0, 2,129.0) | 1,947.0<br>(1,542.5, 2,905.0) | 0.0012 |
|  | Overall<br>N = 2,581<sup>1</sup> | Non-severe<br>N = 2,484<sup>1</sup> | Severe<br>N = 97<sup>1</sup> | p-value<sup>2</sup> | Overall<br>N = 824<sup>1</sup> | Non-severe<br>N = 788<sup>1</sup> | Severe<br>N = 36<sup>1</sup> | p-value<sup>2</sup> |
| sTREM1 (pg/ml)* | 227.0<br>(165.0, 333.0) | 224.0<br>(163.0, 322.0) | 415.0<br>(267.0, 615.0) | < 0.0001 | 238.0<br>(182.0, 331.0) | 235.0<br>(181.0, 325.0) | 359.0<br>(279.5, 453.0) | < 0.0001 |
| suPAR (ng/ml)* | 4.1<br>(3.3, 5.0) | 4.0<br>(3.3, 5.0) | 5.1<br>(3.8, 7.9) | < 0.0001 | 4.6<br>(3.7, 5.8) | 4.5<br>(3.7, 5.7) | 5.6<br>(4.6, 7.2) | 0.0001 |
| **Other laboratory biomarkers** |  |  |  |  |  |  |  |  |
| Lactate (mmol/l)* | 1.0<br>(0.7, 1.3) | 1.0<br>(0.7, 1.3) | 1.1<br>(0.7, 1.8) | 0.0060 | 1.2<br>(0.9, 1.6) | 1.1<br>(0.8, 1.6) | 1.8<br>(1.3, 2.8) | < 0.0001 |
| Glucose (mmol/l)* | 5.4<br>(4.8, 6.2) | 5.4<br>(4.8, 6.1) | 6.0<br>(5.1, 7.1) | < 0.0001 | 5.5<br>(4.9, 6.4) | 5.5<br>(4.9, 6.4) | 5.9<br>(5.0, 7.1) | 0.14 |
| Hb (g/dL)* | 11.5<br>(10.6, 12.4) | 11.5<br>(10.6, 12.4) | 10.9<br>(9.5, 12.2) | 0.0019 | 10.8<br>(9.9, 11.6) | 10.8<br>(9.9, 11.6) | 10.5<br>(9.4, 11.5) | 0.23 |
| <sup>1</sup>Median (Q1, Q3); n (%); <sup>2</sup>Wilcoxon rank sum test; Pearson's Chi-squared test; Fisher's exact test |  |  |  |  |  |  |  |  |

<sup>a</sup>Overnight admission in last six months; <sup>b</sup>Calculated in children aged 3-60 months (R package: *zscorer*);<sup>68</sup> <sup>c</sup>Receipt of any treatment for the current illness in the community prior to presentation at the study site (of these, none had been admitted and 193 had received parenteral treatment); <sup>d</sup>Travel time to the study site ≤ 1 hour; <sup>e</sup>Assessed using the Alert Voice Pain Unresponsive (AVPU) scale. CRT = capillary refill time; LRTI = lower respiratory tract infection; URTI = upper respiratory tract infection.

### Primary outcome: progression to severe febrile illness

Overall, 133 children met the primary outcome (defined as death or organ support within two days of enrolment; 133/3,405; 3.9%): 22 deaths and 111 survivors who required organ support (derivation cohort = 97/2,581; 3.8% and validation cohort = 36/824; 4.4%). Weighted outcome prevalence was 0.34% (95% confidence interval [CI] = 0.28-0.41), which was similar across derivation (0.36%; 95% CI 0.29-0.45) and validation (0.30%; 95% CI 0.20-0.42) cohorts.

Among the candidate clinical predictors (Table S6; appendix p10), association with the outcome was similar across derivation and validation cohorts (Table 1), although a weaker association was observed for altered mental state, prolonged capillary refill time, and presence of WHO danger signs in the validation cohort. Most candidate biomarker predictors were associated with the outcome across derivation and validation cohorts, apart from CRP, IL-1ra, IL-6, IL-10, glucose, and haemoglobin which were not associated with the outcome in the validation cohort.

### Clinical prediction models

When each of the 17 biomarkers were included in turn alongside the candidate clinical predictors, 11 were retained following backward stepwise selection in the derivation dataset. Similar results were obtained with and without the inclusion of SpO_2_ as a candidate predictor (Table S11-12; appendix p17-18). In a sensitivity analysis excluding the northern Viet Nam site (see Methods and Table S13; appendix p19), a biomarker was retained in seven of the models. Across all analyses, the *clinical-biomarker models* containing soluble TREM1 (sTREM1) consistently demonstrated highest discrimination. Thus, four models were taken forward for validation: the *clinical model* (clinical parameters only), the *pulse oximetry model* (clinical parameters plus SpO_2_), the *sTREM1 model* (clinical parameters plus sTREM1), and the *combined model* (clinical parameters plus SpO_2_ and sTREM1).

Heart rate, respiratory rate, prostration, and intractable vomiting were consistently selected across all models (n=4), altered mental state was selected in two models, while prolonged capillary refill time and convulsions were selected in one model each (Table 2). Model equations and odds ratios are reported in the appendix (Tables S14; p20-21).

**TABLE 2.** Selected variables, discrimination, and calibration of the clinical prediction models in the validation cohort.

| MODEL | CONSTITUENT VARIABLES | DISCRIMINATION | CALIBRATION |  |
| --- | --- | --- | --- | --- |
|  |  | AUC (95% CI) | INTERCEPT (95% CI) | SLOPE (95% CI) |
| **Clinical** | Heart rate (bpm)<br>Respiratory rate (bpm)<br>Prostration<br>Intractable vomiting<br>Altered mental state<br>Prolonged capillary refill time | 0.95 (0.92-0.97) | 0.31 (-0.08-0.71) | 1.46 (1.19-1.73) |
| **Pulse oximetry** | Heart rate (bpm)<br>Respiratory rate (bpm)<br>Prostration<br>Intractable vomiting<br>Convulsions<br>Oxygen saturation (%) | 0.97 (0.96-0.99) | 0.60 (0.21-0.99) | 1.74 (1.41-2.06) |
| **sTREM1** | Heart rate (bpm)<br>Respiratory rate (bpm)<br>Prostration<br>Intractable vomiting<br>Altered mental state<br>sTREM1 (pg/ml) | 0.96 (0.93-0.98) | 0.18 (-0.26-0.62) | 1.33 (1.04-1.63) |
| **Combined** | Heart rate (bpm)<br>Respiratory rate (bpm)<br>Prostration<br>Intractable vomiting<br>Oxygen saturation (%)<br>sTREM1 (pg/ml) | 0.98 (0.96-0.99) | 0.48 (0.05-0.91) | 1.58 (1.21-1.96) |

Variables selected in the different clinical prediction models (*clinical model* = clinical parameters only; *pulse oximetry model* = clinical parameters plus SpO<sub>2</sub>; *sTREM1 model* = clinical parameters plus sTREM1; *combined model* = clinical parameters plus SpO<sub>2</sub> and sTREM1). Performance of each model in the validation cohort is summarised using their weighted discrimination and calibration.

Discrimination and calibration of the models in the validation cohort are presented (Table 2). Discrimination ranged from a weighted area under the receiver operating characteristic curve (AUC) of 0.95 to 0.98. Calibration was best at lower predicted probabilities, with some underestimation of risk in individuals at the highest risk of severe disease.

### Clinical utility

Clinical utility of the models for triage of febrile children was assessed within a hypothetical traffic light framework whereby any child with a predicted probability of developing severe disease < 0.5% (the rule-out threshold) is discharged (green), those with a predicted probability > 2% (the rule-in threshold) are referred for higher-level care (red), and those with predicted probabilities between 0.5-2% are monitored (amber), for example, at the rural clinic or via telephone or outreach follow-up.

Within this framework the *clinical model* recommended discharge in 90.6%, monitoring in 8.4%, and referral in 1.0% of presentations (Figure 1); one in 10 referred children progressed to develop severe disease (positive likelihood ratio [PLR] 37.5; 95% CI 14.3-107.6). The *clinical model* failed to identify 25% of children who progressed to severe disease, in whom discharge was recommended (sensitivity 0.75; 95% CI 0.62-0.86). Compared to the *clinical model*, the *pulse oximetry model* recommended fewer referrals (0.3% vs. 1.0%) and was better able to rule-in children at risk of disease progression with one in three referrals developing severe disease (PLR 150.7; 95% CI 84.3-249.0). It also recommended more children for discharge (94.7% vs. 90.6%) while demonstrating superior rule-out performance (sensitivity 0.89; 95% CI 0.79-0.97). In addition, fewer children were recommended for monitoring (5.0% vs. 8.4%).

**FIGURE 1.**
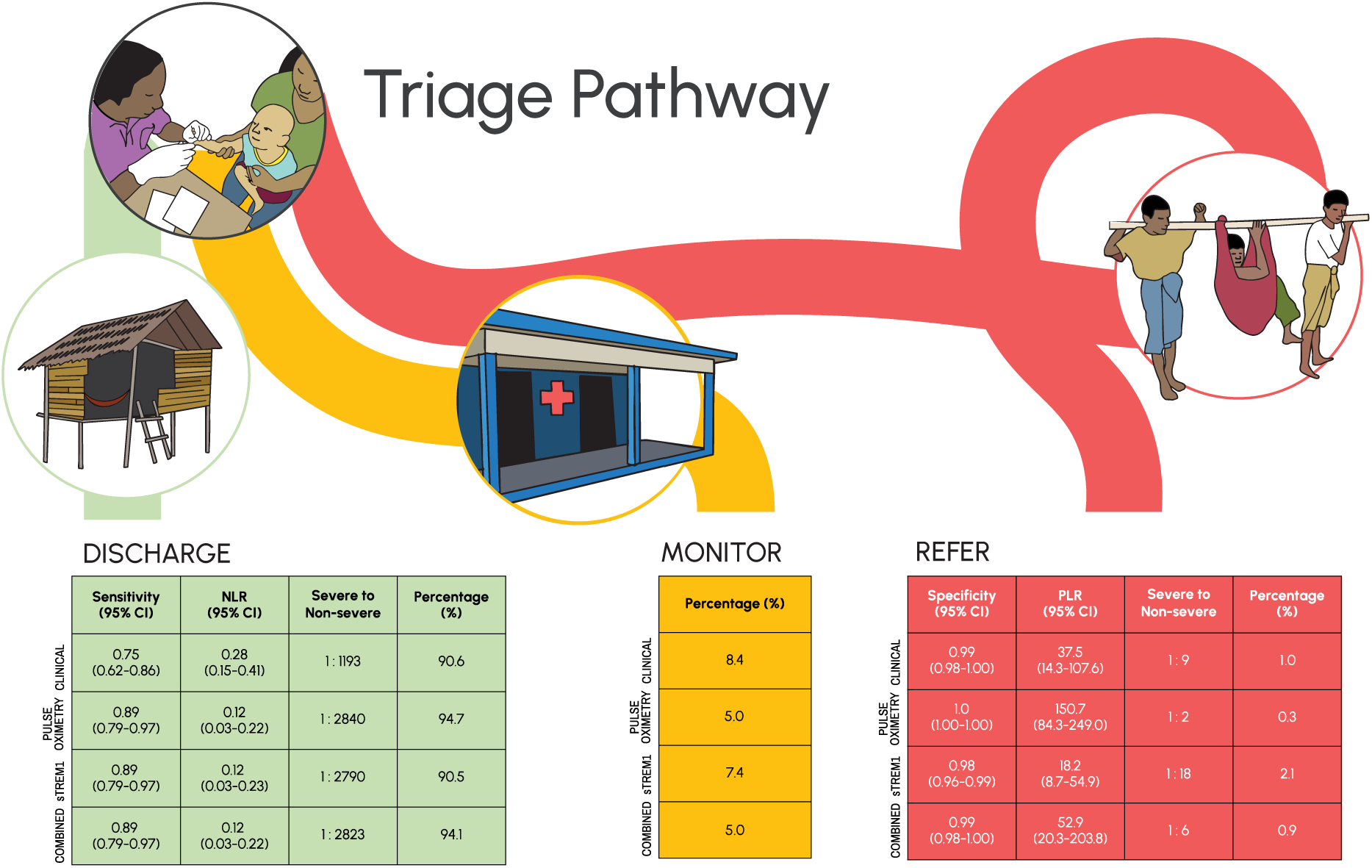
Utility of the clinical prediction models within a hypothetical traffic light triage framework. Among patients recommended for discharge (green; predicted probability of severe disease [pp] < 0.5%), the sensitivity and NLR indicate the ability of the model to rule-out severe disease. The percentage of patients recommended for monitoring (amber; pp 0.5-2%) indicates the potential burden on the health system created by the resources required for observation or proactive safety-netting. Among patients recommended for referral (red; pp > 0.5%), the specificity and PLR indicate the ability of the model to rule-in severe disease, and the percentage of patients indicates the potential burden on the health system created by the resources required for referral. *Clinical model* = clinical parameters only; *pulse oximetry model* = clinical parameters plus SpO_2_; *sTREM1 model* = clinical parameters plus sTREM1; *combined model* = clinical parameters plus SpO_2_ and sTREM1. NLR = negative likelihood ratio; PLR = positive likelihood ratio; pp = predicted probability.

Compared to the *clinical model*, the *sTREM1 model* improved rule-out performance to a similar extent as the *pulse oximetry* model (sensitivity 0.89; 95% CI 0.79-0.97) but recommended more referrals (2.1% vs 1.0%) translating to poorer rule-in performance (PLR 18.2 vs. 37.5). The *combined model* (including both SpO_2_ and sTREM1) did not offer advantage over the *pulse oximetry model*, neither in terms of discharge (rule-out) or referral (rule-in) performance nor the overall number of children recommended for monitoring or referral.

At the outcome prevalence observed in the validation cohort (0.30%), compared to using the *clinical model*, either the *pulse oximetry model* or *sTREM1 model* would prevent one additional child who would progress to life-threatening infection being discharged for every ∼2,300 children tested (number needed to test [NNT] = 2,300). For the *pulse oximetry model* these gains would be achieved in addition to a three-fold reduction in the referral rate (0.3% vs 1.0%), whilst for the *sTREM1 model* the gains would be partially offset by twice the number of referrals (2.1% vs 1.0%) compared to the *clinical model*.

All four models outperformed WHO danger signs in terms of discharge or rule-out (sensitivity 0.56; 95% CI 0.42-0.69) and referral or rule-in (specificity 0.83; 95% CI 0.78-0.87) performance. Compared to WHO danger signs, the prediction models could prevent one additional child who would progress to life-threatening infection being discharged for every 1,000 (*pulse oximetry*, *sTREM1*, or *combined models*) or 1,750 (*clinical model)* children tested (NNT), whilst simultaneously substantially reducing referral rates from 17.0% to 0.3-2.1%. As WHO danger signs provide a binary classification (refer or not), any benefits gained through use of the prediction models would be partially offset by the costs of monitoring up to 8.4% of all presentations.

### Categorical outcome scale

All four models predicted increasing probability of progression to severe febrile illness at higher levels of the categorical outcome scale (Table S4; appendix p7) in the validation cohort (Figure 2), suggesting they were able to provide discrimination across the severity spectrum, rather than only identify the most severely unwell patients.

**FIGURE 2.**
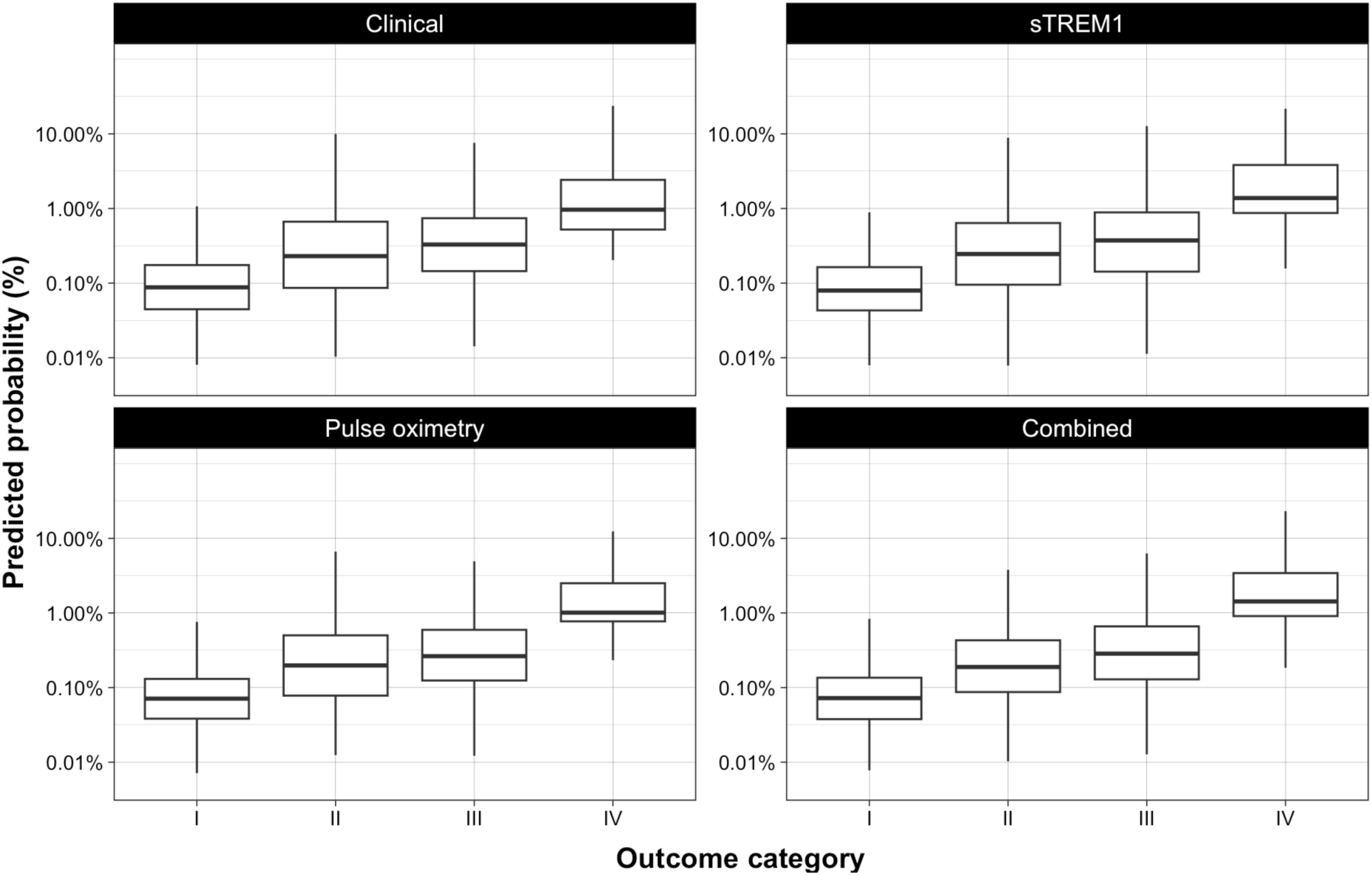
Predicted probability of severe febrile illness across the categorical outcome scale for each prediction model in the validation cohort. *Clinical model* = clinical parameters only; *pulse oximetry model* = clinical parameters plus SpO_2_; *sTREM1 model* = clinical parameters plus sTREM1; *combined model* = clinical parameters plus SpO_2_ and sTREM1. Category IV = death or organ support within two days of enrolment; Category III = admission to any health facility for > two nights between enrolment and day 28; Category II = admitted to any health facility for ≤ two nights between enrolment and day 28; Category I = managed as an outpatient and recovered by day 28.

Using the categorical outcome scale to assess triage correctness, indicated that improvement in sensitivity of the other models compared to the *clinical model* was driven by increases in both referrals and monitoring of the most severe (category IV) patients (Figure 3). The reduction in referrals achieved by the *pulse oximetry model* was a consequence of both more discharges and fewer cases being recommended for monitoring among the least unwell children (category I).

**FIGURE 3.**
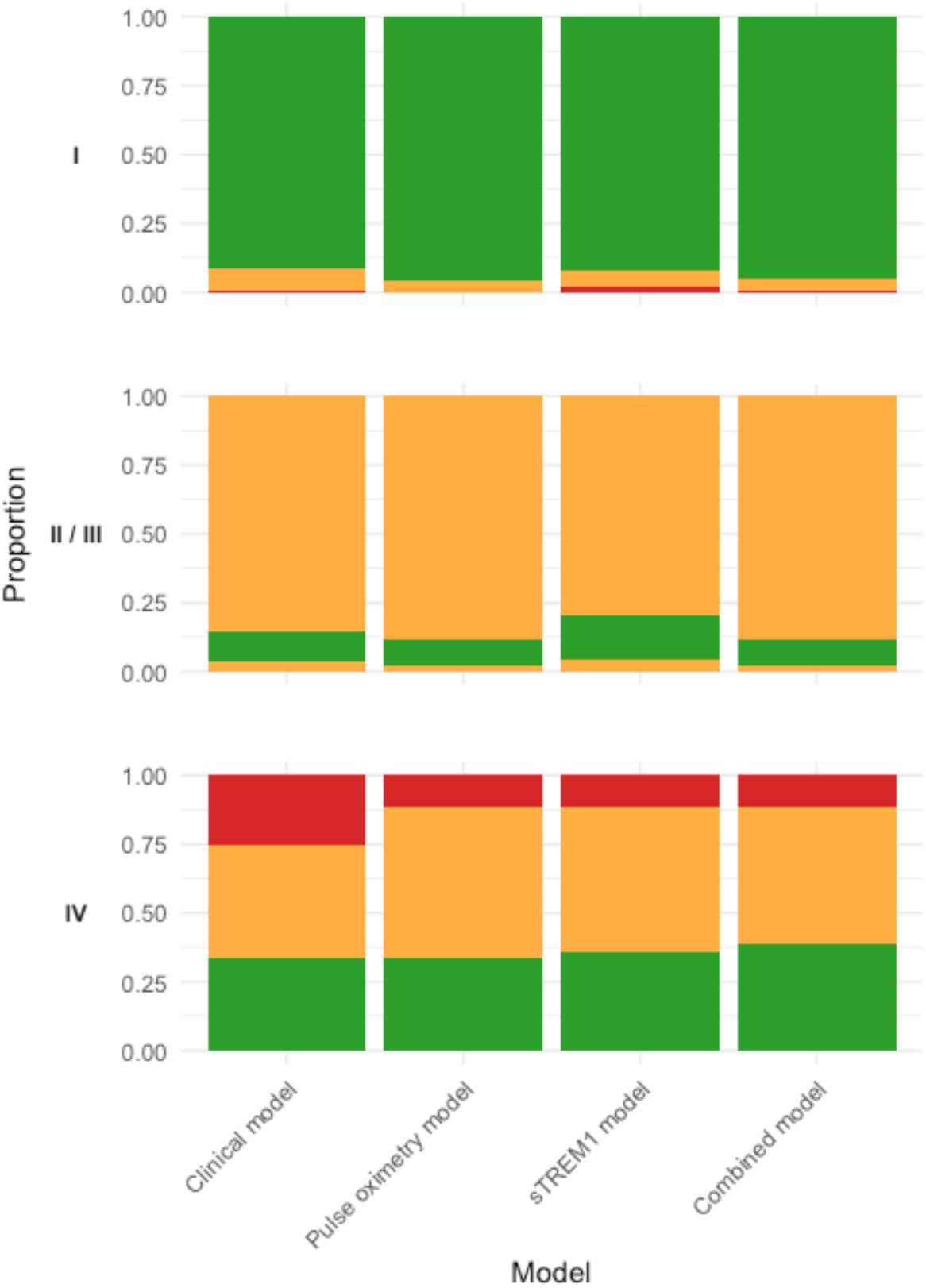
Proportional distribution of triage groups by patient category for each prediction model. Proportional disposition of patients in each categorical outcome category is colour-coded to reflect triage appropriateness. Category I (least severe): discharged patients (pp < 0.5%) = green; observed patients (pp 0.5-2%) = amber; referred patients (pp > 2%) = red. Category II and III (moderately severe): discharged patients = amber; observed patients = green; referred patients = amber. Category IV (most severe): discharged patients = red; observed patients = amber; referred patients = green. Within each outcome category (horizontal facet), patients are ordered as discharged, observed, and referred, from top to bottom. *Clinical model* = clinical parameters only; *pulse oximetry model* = clinical parameters plus SpO_2_; *sTREM1 model* = clinical parameters plus sTREM1; *combined model* = clinical parameters plus SpO_2_ and sTREM1. Category IV = death or organ support within two days of enrolment; Category III = admission to any health facility for > two nights between enrolment and day 28; Category II = admitted to any health facility for ≤ two nights between enrolment and day 28; Category I = managed as an outpatient and recovered by day 28. pp = predicted probability of severe febrile illness.

### Missed cases of severe febrile illness

Among the 133 participants that progressed to severe febrile illness, the same children were missed by all four models in both the derivation and validation cohort (Figure 4; Table S8; appendix p11-12). Further, the additional participants identified by the *pulse oximetry*, *sTREM1*, and *combined models* were consistent across these three models.

**FIGURE 4.**
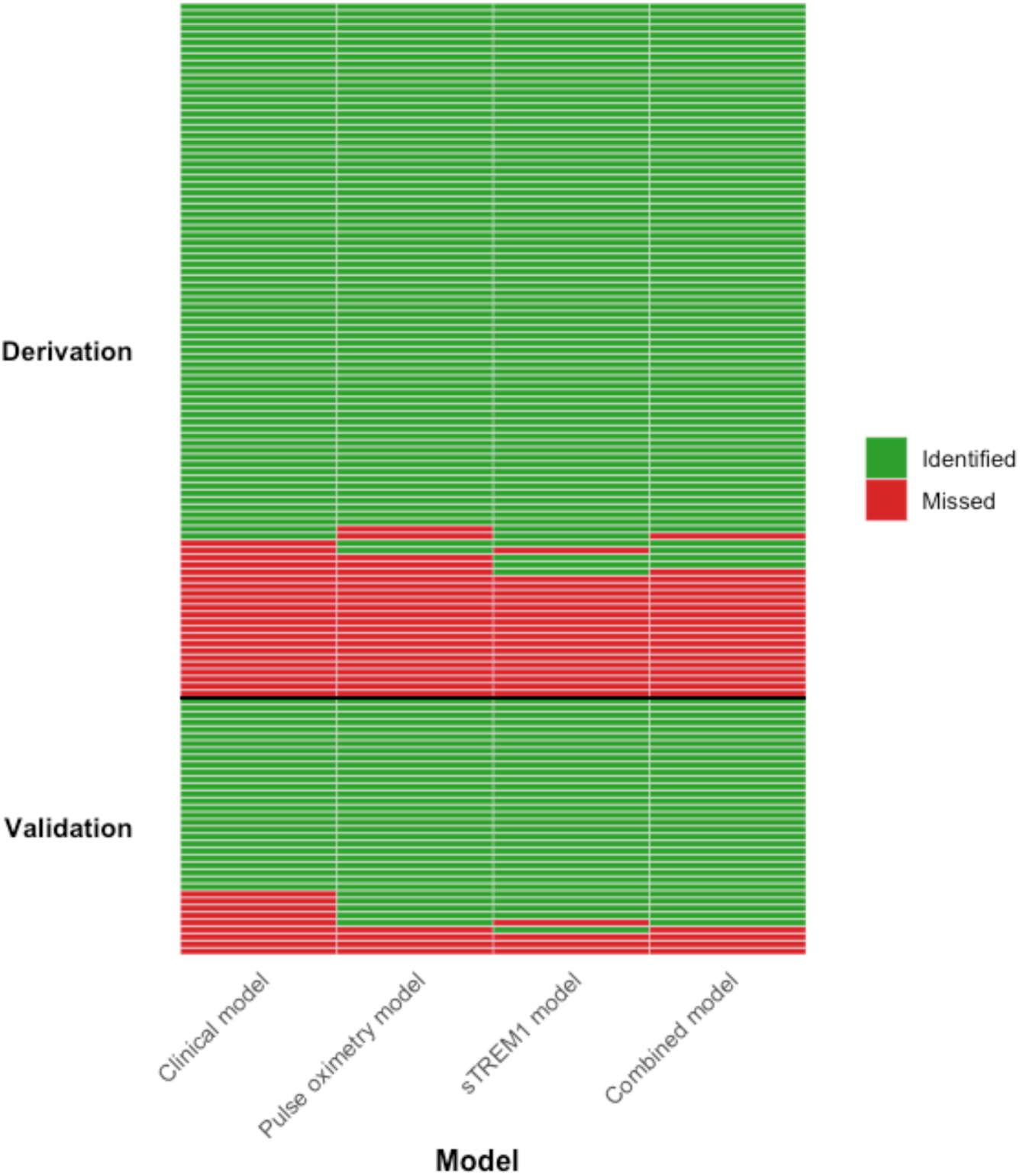
Identification of participants who progressed to severe febrile illness for each of the models, split by derivation and validation cohort. Classification for each of the 133 participants who progressed to develop severe febrile illness according to whether they were identified (green; pp > 2%) or missed (red; pp ≤ 2%) by each of the models. *Clinical model* = clinical parameters only; *pulse oximetry model* = clinical parameters plus SpO_2_; *sTREM1 model* = clinical parameters plus sTREM1; *combined model* = clinical parameters plus SpO_2_ and sTREM1. pp = predicted probability of severe febrile illness.

Older children, males, those with a shorter illness prior to presentation, and those without pneumonia, were more likely to be missed by the models (Table S15; appendix p22-23).

Baseline biomarker concentrations were typically less deranged in children the models failed to identify, apart from ANG-1, CRP, IL-6, and IP-10 concentrations, which were more abnormal in children who progressed to severe disease but were missed by the models.

The models were more likely to identify children who progressed to severe illness quickly. Extending the definition of severe febrile illness to include all cases that developed in the 28 days following enrolment, identified an additional 10 participants (two deaths and eight survivors who required organ support). Among the 42 children who progressed to develop severe febrile illness more than 24 hours after enrolment, 12 (28.6%) were missed by the *clinical model*, whilst the *pulse oximetry* and *sTREM1 models* missed 9 (21.4%) and 10 (23.8%) participants respectively.

### Cost-effectiveness

All four models were predicted to dominate (i.e. were cost-saving and more effective than) WHO danger signs. Compared to the *clinical model*, the *pulse oximetry*, *sTREM1* and *combined models* were all predicted to be cost-effective using both cost-effectiveness thresholds (CETs): $2,551/DALY averted and $459/DALY averted (Table 3).^48–50^ Cost-effectiveness for the *combined* and *pulse oximetry models* was predicted to improve in contexts with higher referral costs, as a result of their better specificity compared to the *clinical model*. Although specificity for the *sTREM1 model* was inferior to the *clinical model*, it was predicted to remain cost-effective up to a referral cost of ∼$600 per patient, even when using the conservative CET (Table S16; appendix p24). Neither the *sTREM1* nor *combined models* were predicted to be more cost-effective than the *pulse oximetry model*, at either of the analysed CETs.

**TABLE 3.**
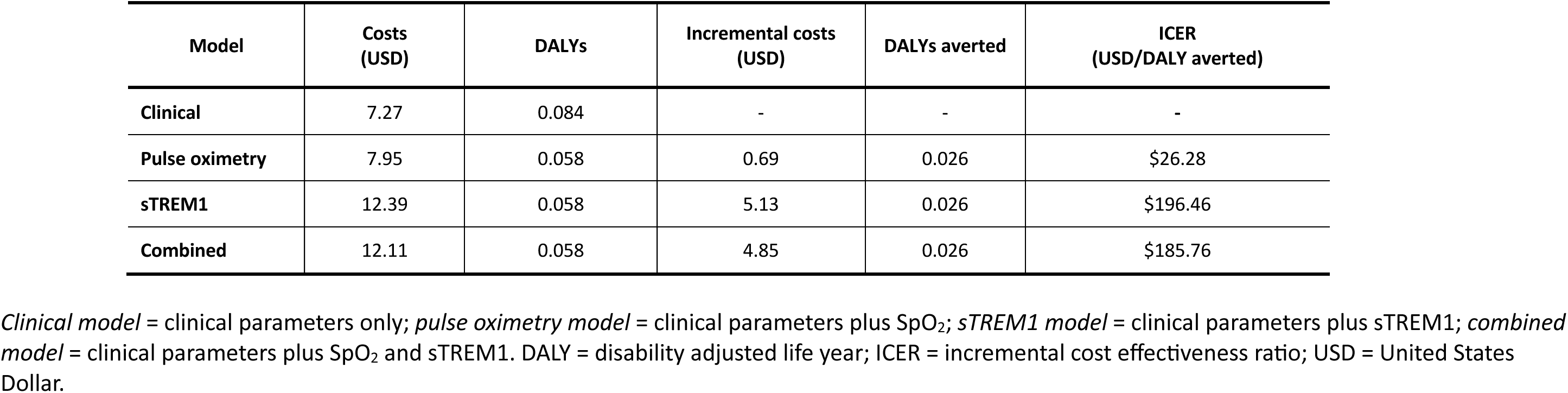
Cost effectiveness of the *pulse oximetry*, *sTREM1*, and *combined models* compared to the *clinical model*.

| Model | Costs (USD) | DALYs | Incremental costs (USD) | DALYs averted | ICER (USD/DALY averted) |
| --- | --- | --- | --- | --- | --- |
| Clinical | 7.27 | 0.084 | - | - | - |
| Pulse oximetry | 7.95 | 0.058 | 0.69 | 0.026 | \$26.28 |
| sTREM1 | 12.39 | 0.058 | 5.13 | 0.026 | \$196.46 |
| Combined | 12.11 | 0.058 | 4.85 | 0.026 | \$185.76 |

*Clinical model* = clinical parameters only; *pulse oximetry model* = clinical parameters plus SpO<sub>2</sub>; *sTREM1 model* = clinical parameters plus sTREM1; *combined model* = clinical parameters plus SpO<sub>2</sub> and sTREM1. DALY = disability adjusted life year; ICER = incremental cost effectiveness ratio; USD = United States Dollar.

## DISCUSSION

This multi-country prospective study, conducted in seven locations across Asia, developed and validated clinical prediction models which outperform the current standard of care for triage of febrile children in resource-constrained community settings. While all models were superior to WHO danger signs, integration of pulse oximetry or host biomarker testing alongside measurement of simple clinical parameters was required to adequately rule-out impending critical illness, a key consideration for effective risk stratification at the community level. Importantly, our analyses indicate that both strategies are likely to be cost-effective across a range of CETs, including in conflict-affected settings and remote contexts, where referral costs are often high.

Our findings align with previous studies which identified the prognostic potential of specific clinical parameters in paediatric febrile illness,^35,52–57^ and build on emerging evidence supporting hypoxaemia and host biomarkers, such as sTREM1, as indicators of poor outcome.^17,20,25,58,59^ The superior performance of the prediction models compared to previous univariate analyses underscores the importance of a multivariate approach to risk stratification,^30,35,60^ whereby combinations of multiple “nearly” signs can prompt action even if no “absolute” red flags are present.^61^

The *pulse oximetry model* outperformed the *sTREM1 model* and is likely to be the preferred option in many settings. Its adoption would be particularly suited to primary health centres and rural clinics with access to safe storage, secure supply chains, supplemental oxygen therapy, and staff trained to manage other conditions for which a pulse oximeter may provide benefit. By contrast, for lesser-trained community health workers, who typically have a limited scope of practice and are accustomed to screening all febrile children for malaria, a triage tool incorporating host biomarker testing may be more practical, particularly if multiplexed with a rapid diagnostic test for malaria.^31,62^

It is important to understand which patients at risk of severe illness may be missed by a particular triage approach. Our models appear to perform best in younger children with pneumonia – a group that carries a disproportionate share of the disease burden, indicating opportunity for substantial public health impact.^1^ Conversely, children presenting earlier in their illness or deteriorating later were more likely to be missed, suggesting a need for adjusted approaches to identify these children. Prior work has shown that host biomarkers may be particularly valuable for identifying children whose illness severity is not clinically evident at presentation and who are at risk for delayed clinical deterioration.^30,63^ In our study, both the *pulse oximetry* and *sTREM1 models* improved identification of such children relative to the *clinical model*, though small event numbers in this subgroup mean that further work is warranted.

To minimise outcome misclassification, we applied a stringent definition of severe illness – restricted to children that died or received organ support within two days of enrolment – to develop the prediction models. Consequently, the reported NNTs and severe to non-severe referral ratios represent conservative estimates, emphasising model performance in identifying children who progressed to the most severe illness. In practice, however, a substantially larger proportion of children would merit hospital referral. Secondary analyses utilising the categorical outcome indicate that the models provide discrimination across the severity spectrum, and further work is underway to quantify this performance.

To our knowledge, this is the first multi-country study to develop and validate risk stratification tools for febrile children presenting directly from the community. We overcame weaknesses of previous studies by adhering to best-practice methodology for clinical prediction model building; pre-specifying candidate predictors, avoiding overfitting, and utilising a held-out geographic validation dataset to assess model performance.^51^ Other strengths include eligibility criteria and a recruitment strategy which ensured a study population and outcome prevalence reflective of community care settings.^46,47^ Our analytical framework prioritised diagnostic accuracy, clinical utility, and cost-effectiveness, over summary measures of model performance such as the AUC.^64^

Several limitations must be discussed. Despite efforts to approximate community settings, our study population were recruited at rural hospital outpatient departments and differences between these patients and those seeking care at primary care facilities may remain. The relatively short interval to developing severe disease may indicate a higher baseline severity than is typical in some community care settings. Use of trained research assistants may have inflated the apparent performance of clinical parameters – in routine care settings where inter-observer variability is high, it may be that incremental benefit of pulse oximetry and host biomarker testing could be even greater.^13^

Model calibration was sub-optimal, likely due to differences in the prevalence of key clinical predictors – such as intractable vomiting and convulsions – between the derivation and validation cohorts. These discrepancies may reflect the well-documented inter-observer variability in assessing WHO danger signs, suggesting that excluding these variables could improve model performance.^13^ While standardised assessments might mitigate this issue, in our study, trained research assistants used precise definitions; under routine care, variability is likely to be greater. Miscalibration primarily involved underestimation of risk among the highest-risk individuals – critically, the models still advised referral in these cases.

Nonetheless, further external validation is needed. We have published our full models to encourage independent validation.

In our cohort, malaria was uncommon, consistent with the evolving epidemiology of febrile illness across Asia.^65^ Circulating markers of immune and endothelial activation, such as sTREM1, appear to capture convergent pathways of host response and enable risk stratification of paediatric fever syndromes irrespective of underlying aetiology, including both malarial and non-malarial infections.^25,38,66^ Similarly, hypoxaemia has emerged as a robust predictor of adverse outcome across a spectrum of common childhood infections.^17,20^ Nonetheless, evaluation of model performance in malaria-endemic regions will be essential to establish generalisability and clinical utility.

Among the *clinical-biomarker models*, we selected the *sTREM1 model* for external validation because it consistently performed best across development and sensitivity analyses, retained face validity from prior studies,^25,27,30,59^ and is under active development as a point-of-care assay.^67^ Nonetheless, optimism-adjusted performance was similar across several candidate biomarkers, and other *clinical-biomarker models* may prove equally or more effective. Limiting models to a single biomarker reduced overfitting and improves feasibility for application on the field, but future work should examine combinations of biomarkers, balancing potential gains in accuracy against increased cost and complexity. Including biomarkers found to be elevated in children who progressed to severe illness but missed by the current models (for example, ANG-1, CRP, IL-6, or IP-10) may be a fruitful approach.

We envisage these models being applied selectively to children for whom referral decisions are borderline and no overt signs of severe illness are evident at presentation. The exact ‘red flags’ triggering referral will differ across settings and are influenced by health worker capacity, safety netting options, treatments available in the community, opportunity costs of referral, and the quality of care at the higher-level facility. In our cohort, some children may be considered as presenting with features mandating referral and would not be appropriate candidates for model-based triage in all settings. Careful specification of referral criteria and further refinement and contextualisation of the models will be essential prior to evaluation in clinical trials or integration into routine care.

The traffic light framework we used to illustrate clinical utility reflects the reality that in most settings some sort of safety-netting is usually possible.^61^ Decision thresholds were selected pragmatically based on field experience but will require adaptation to local contexts. Future work should evaluate how different decision thresholds influence model performance and health system burden.

In summary, we have developed and validated clinical prediction models that substantially improve risk stratification of febrile children compared with current practice. Incorporation of pulse oximetry or sTREM1 measurements alongside simple clinical parameters achieves triage performance that may be acceptable to patients, providers, and policy makers. Both approaches appear cost effective across diverse contexts, including those with high referral costs. Further external validation and interventional trials will be essential to confirm clinical impact and support policy adoption.

## AUTHOR CONTRIBUTIONS

AC, CK, RAA, PT, MM, EAA, EA, RP-S, MR-G, YL, and SB conceptualised the study. AC, DTVA, KSC, SK, PNTN, SR, SV, PHP, DM, BTL, and EA acquired the data. AC, RM, MY, NW, and MR-G curated the data. MYA, JT, and MR-G performed the laboratory assays. AC, CK, RM, and RP-S did the formal analysis. YL and SB acquired funding. AC wrote the original draft of the manuscript. All authors reviewed and edited the manuscript. AC, CK, RM, MR-G, YL, and SB verified the underlying data. AC, CK, YL, and SB had full access to all the data in the study and had final responsibility for the decision to submit for publication.

## Data Availability

De-identified, individual participant data from this study will be available to researchers whose proposed purpose of use is approved by the data access committees at MSF and MORU. Enquiries or requests for data can be sent to. Researchers interested in accessing biobanked samples should contact the corresponding author, who will coordinate with the Spot Sepsis Sample Use Committee.

## ACKNOWLEDGEMENTS

We are extremely grateful to all the participants and their caregivers, and the laboratory and research staff at the participating sites and MORU laboratories in Bangkok for specimen processing, data management, and study monitoring. We acknowledge the support of bioMérieux (Marcy-l’Étoile, France), which provided laboratory consumables and equipment for some of the respiratory specimen assays. We also acknowledge the valuable support and guidance of Nicholas J White (MORU) at key points during the study.

## OPEN ACCESS

This research was co-funded by MSF and Wellcome [219644/Z/19/Z]. For the purpose of open access, the authors have applied a CC BY public copyright license to any Author Accepted Manuscript version arising from this submission.

## COMPETING INTERESTS

We declare no competing interests.

## Notes

### Competing Interest Statement

The authors have declared no competing interest.

### Clinical Protocols

https://bmjopen.bmj.com/content/11/1/e045826

### Funding Statement

The study was co-funded by Medecins Sans Frontieres, Spain (MSF) and Wellcome. MSF maintained a sponsor-investigator role for the study. Wellcome had no role in study design, data collection, data analysis, data interpretation, writing of the report, or decision to submit for publication.

### Author Declarations

Medecins Sans Frontieres Ethical Review Board; Oxford Tropical Medicine Research Committee; International Centre for Diarrhoeal Disease Research; Angkor Hospital for Children Research Committee; National Ethics Committee for Health Research, Cambodia; Medical and Health Research Ethics Committee, Indonesia; National Ethics Committee for Health Research, Lao PDR; University of Medicine and Pharmacy at Ho Chi Minh City, Viet Nam; Ethics Committee for Biomedical Research, Viet Nam

