## Appendix for "Referral of febrile children in resource-constrained community settings in Asia (Spot Sepsis) – a multi-country, prospective, cohort study"

**SUPPLEMENTARY APPENDIX**

1. **Details of the study sites.**

Sites located outside major urban cities were proactively approached. As part of the shortlisting process, sites were asked to indicate the proportion of their patient population residing in a rural location and the proportion of children using the hospital as a first point of contact with the formal healthcare sector. Where possible, routinely collected data were used to inform site selection. If these were not available, local clinicians and hospital administrators were asked to provide their best estimate.

Due to the disruptions caused by the Covid-19 pandemic, site activation was delayed and recruitment proceeded slower than anticipated at all sites. After 18 months of recruitment, a decision was taken to identify an additional site in order to boost recruitment, ensure sufficient outcome events, and safeguard viability of the study. The second Viet Nam site was subsequently identified and brought online in December 2021, acknowledging that it departed from the desired rural site target profile established at the outset of the study. A sensitivity analysis excluding this site was prespecified in order to address this.

**Bangladesh**

*Recruitment period: 18/03/2021 – 27/04/2022*

Goyalmara Mother and Child Hospital is a non-governmental hospital managed by Médecins Sans Frontières located in Cox’s Bazaar, Chattogram Division, in eastern Bangladesh, which predominantly provides health services to the forcibly displaced Rohingya refugee population. The hospital provides primary and secondary care, with approximately 20,000 outpatient attendances and 4,000 inpatient admissions annually. The 12-bed high-dependency unit provides non-invasive ventilation and inotropic therapy. There is a basic on-site diagnostic laboratory.

**Cambodia**

*Recruitment period: 05/03/2020 – 24/02/2022*

Angkor Hospital for Children is a non-governmental paediatric hospital located in Siem Reap province, northern Cambodia. The hospital provides primary-to-tertiary care, with approximately 80,000 outpatient attendances and 3,000 inpatient admissions annually. The 14-bed intensive care unit provides non-invasive ventilation, mechanical ventilation, inotropic therapy, and peritoneal dialysis. There is an on-site diagnostic microbiology laboratory (ISO15189 accredited since 23 November 2023).

**Indonesia**

*Recruitment period: 22/03/2021 – 22/04/2022*

Rumah Sakit Umum Daerah Wates is a government district hospital located in Yogyakarta province, Indonesia. The hospital provides primary and secondary care, with approximately 40,000 outpatient attendances and 4,000 inpatient admissions annually. The 8-bed intensive care unit provides non-invasive ventilation, mechanical ventilation, inotropic therapy, and renal replacement therapy. Basic microscopy is available on-site. Culture-based microbiology is available via nearby private laboratories.

**Laos 1**

*Recruitment period: 10/09/2020 – 30/08/2021*

Salavan Provincial Hospital is the government provincial hospital for Salavan, a predominantly rural province in southern Laos. The hospital provides primary-to-tertiary care, with approximately 65,000 outpatient attendances and 12,000 inpatient admissions annually. The 3-bed paediatric intensive care unit provides non-invasive ventilation, mechanical ventilation, and inotropic therapy. There is an on-site diagnostic microbiology laboratory.

**Laos 2**

*Recruitment period: 21/01/2021 – 26/08/2021*

Savannakhet Provincial Hospital is the government provincial hospital for Savannakhet, a predominantly rural province in southern Laos. The hospital provides primary-to-tertiary care, with approximately 7,000 outpatient attendances and 3,000 inpatient admissions annually. The 22-bed intensive care unit provides non-invasive ventilation, mechanical ventilation, and inotropic therapy. There is an on-site diagnostic microbiology laboratory.

**Viet Nam 1**

*Recruitment period: 10/05/2021 – 28/10/2022*

Dong Nai Children's Hospital is the government provincial paediatric hospital for Dong Nai province in southern Viet Nam. The hospital provides primary-to-tertiary care, with approximately 80,000 outpatient attendances and 7,000 inpatient admissions annually. The 30-bed intensive care unit provides non-invasive ventilation, mechanical ventilation, inotropic therapy, and renal replacement therapy. There is an on-site diagnostic microbiology laboratory.

**Viet Nam 2**

*Recruitment period: 08/12/2021 – 04/11/2022*

Viet Nam National Children's Hospital is the government national paediatric hospital, located in Ha Noi. The hospital provides primary-to-quaternary care, with approximately 1,200,000 outpatient attendances and 110,000 inpatient admissions annually. The 40-bed intensive care unit provides non-invasive ventilation, mechanical ventilation, inotropic therapy, and renal replacement therapy. There is an on-site diagnostic microbiology laboratory.

### 2. Table S1: Methodology for collection of clinical parameters.

| Variable | Methodology |
|---|---|
| Respiratory rate | Manual count for 60 seconds using clicker counter and timer |
| Heart rate | Massimo Rad-5v pulse oximeter with paediatric and neonatal probes |
| Oxygen saturation | Massimo Rad-5v pulse oximeter with paediatric and neonatal probes |
| Axillary temperature | Digital thermometer: operating range 32.0-42.9°C; accuracy ± 0.1°C |
| Capillary refill time | Pressure applied to sternum for 5 seconds |
| Length / Height | *Médecins Sans Frontières* height and length board |
| Weight | Seca 877 scale with mother-and-child function; accuracy ± 50g |
| Mid-upper arm circumference | *Médecins Sans Frontières* traffic light MUAC tape |
| Mental state | Alert Voice Pain Unresponsive (AVPU) scale |
| WHO danger signs | WHO IMCI Distance Learning course: https://iris.who.int/handle/10665/104772 |

### 3. Table S2: Existing evidence supporting selection of endothelial and immune activation markers.

| Biomarker | Overview of supportive data |
|---|---|
| **Endothelial activation** | |
| **ANG-1 and -2** | Supportive data from Asia/SSA/Europe in children/adults, that increases in ANG-2, decreases in ANG-1, and/or the ANG-2:1 ratio predicts mortality in pneumonia, malaria, SBI, and all-cause febrile illnesses,[1-10] and supplemental oxygen requirement in children with pneumonia in Asia.[11] |
| **sFLT-1** | Supportive data from SSA that increases in sFLT-1 predict mortality in children hospitalised with pneumonia, severe malaria, and all-cause febrile illnesses, and adults with all-cause febrile illnesses.[1,5,6,9,12] |
| **Immune activation** | |
| **CHI3L1** | Supportive data from SSA that increases in CHI3L1 predict mortality in children hospitalised with pneumonia and all-cause febrile illnesses, and adults with all-cause febrile illnesses.[1,5,6] |
| **CRP** | Although there is limited supportive evidence for the use of CRP as a prognostic marker for disease severity,[13] as it is the most widely studied biomarker in the region, and numerous point-of-care tests already exist, further evaluation is warranted. |
| **IL-1ra** | Supportive data that increases in IL-1ra are associated with severity in children with meningococcal disease, adults with SARS-CoV-2 infection, and predict need for longer antibiotic duration in children with febrile lower respiratory tract infections.[14-16] |
| **IL-6** | Supportive data from India that increases in IL-6 are predictive of mortality in children with dengue;[17] in Switzerland, supportive data that increases in IL-6 predict need for longer antibiotic duration in children with febrile lower respiratory tract infection, and disease severity in adults with SARS-CoV-2 infection.[15,18] |
| **IL-8** | Supportive data from India that increases in IL-8 predict mortality in children with dengue;[17] in Mozambique, IL-8 predicted mortality in children with pneumonia;[12] in the UK, supportive data that increases in IL-8 predict disease severity in children with meningococcal disease.[14] |
| **IL-10** | Supportive data from India that increases in IL-10 predict of mortality in children with dengue.[17] |
| **IP-10** | Supportive data from Uganda that increases in IP-10 predict mortality in children hospitalised with severe malaria.[9] |
| **PCT** | Supportive evidence that increases in PCT predict severe illness in hospitalised children with suspected bacterial infections or meningococcal disease.[19,20] |
| **sTNF-R1** | Supportive data from SSA that increases in sTNF-R1 predict mortality in children hospitalised with pneumonia and all-cause febrile illnesses, and adults with all-cause febrile illnesses.[1,5,6] |
| **sTREM1** | Supportive data from SSA that increases in sTREM1 predict mortality in children hospitalised with pneumonia, severe malaria, and all-cause febrile illnesses, and adults with all-cause febrile illnesses;[1,5,6,9,12] in Asia, increased sTREM1 predicted length of stay in infant febrile illness and in-hospital mortality in adults hospitalised with infection and children hospitalised with pneumonia.[7,21,22] |
| **suPAR** | Supportive data from Uganda that increases in suPAR predict mortality in children with malaria;[23] In Europe, elevated suPAR concentrations predicted length of stay in children with pneumonia and mortality in adults hospitalised with sepsis.[24-26] |

### 4. Table S3: Laboratory procedures for biomarker quantification.

Host biomarker concentrations were quantified in EDTA-plasma using the Simple Plex Ella microfluidic platform (ProteinSimple, San Jose, CA, USA) and suPARnostic ELISA (ViroGates, Denmark), or fluoride-oxalate-plasma using LACT2 (Roche Diagnostics, Germany) and GLUC3 (Roche Diagnostics, Germany), according to the manufacturers' protocols. All biomarkers were quantified at the MORU laboratories in Bangkok, Thailand, apart from Indonesian samples, which were quantified at the INA-RESPOND laboratories in Jakarta, Indonesia, with consumables and on-site support provided by the visiting MORU laboratory team.

For the Ella platform, plasma samples were diluted 1:2 (ANG-2, IL-6, IL-8, IL-10, PCT, sTREM1, sFLT-1), 1:10 (ANG-1, CHI3L1, IL-1ra, IP-10, sTNF-R1), or 1:5000 (CRP) in reagent diluent. Analyte concentrations outside the dynamic range of the assay using the initial dilutions were prepared at higher or lower dilutions depending on the raw data relative fluorescent units (RFUs). For suPAR, no dilutions to plasma were performed prior to adding the samples to the pre-coated ELISA plate. Any samples with concentrations above or below the assays' limit of detection were assigned a value of one-third of the highest or lowest limit, respectively, of the limits of quantification. The table below details the proportion of samples assigned a value.

| | **Dynamic Range** (pg/mL) | **Samples outside dynamic range** | **Proportion assigned a value** |
|---|---|---|---|
| ANG-1 | 6.18 - 23,560 | 0 | - |
| ANG-2 | 9.91 - 15,124 | 0 | - |
| CHI3L1 | 6.68 - 25,500 | 2 | 2/3,312 (0.06%) |
| CRP | 32.8 - 50,000 | 5 | 5/3,309 (0.15%) |
| IL-1ra | 7.37 - 4,500 | 1 | 1/3,313 (0.03%) |
| IL-6 | 0.28 - 2,652 | 4 | 4/3,314 (0.12%) |
| IL-8 | 0.19 - 1,804 | 2 | 2/3,314 (0.06%) |
| IL-10 | 0.58 - 2,212 | 2 | 2/3,314 (0.06%) |
| IP-10 | 0.6 - 920 | 0 | - |
| PCT | 1.58 - 15,100 | 0 | - |
| sFLT-1 | 3.05 - 4,650 | 0 | - |
| sTNF-R1 | 0.89 - 3,390 | 0 | - |
| sTREM1 | 4.2 - 40,000 | 1 | 1/3,314 (0.03%) |
| suPAR | 400 - 16,000* | 15 | - |

*Samples outside the dynamic range (>16,000 pg/mL) for suPAR could be estimated correctly up to 25,000 pg/mL and thus new values were not assigned.

### 5. Table S4: Categorical outcome scale.

| Category | Definition |
| --- | --- |
| **I** | Not admitted to any health facility between enrolment and D28 AND recovered by D28 |
| **II** | Admission to any health facility for ≤ 2 nights between enrolment and D28 OR symptoms not resolved by D28 |
| **III** | Admission to any health facility for > 2 nights between enrolment and D28 OR death or organ support between D2 and D28 |
| **IV** | Death or organ support ≤ 2 days after enrolment* |

*Category IV equivalent to the primary outcome. Outcome categories were determined in descending order of severity and were mutually exclusive, i.e. if a participant met the criteria for Category IV, this would be their classification.

6. **Table S5: Derivation of site-specific outpatient weights.**

| | Bangladesh | Cambodia | Laos 1 | Laos 2 | Viet Nam 1 | Viet Nam 2 | Total |
|---|---|---|---|---|---|---|---|
| **Screening week data** | | | | | | | |
| Number of screening weeks | 3 | 4 | 2 | 1 | 2 | - | - |
| Patients screened (A) | 1,556 | 1,872 | 204 | 39 | 899 | - | - |
| Patients eligible (B) | 320 | 346 | 71 | 4 | 180 | - | - |
| Proportion eligible (B/A = C) | 0.21 | 0.19 | 0.35 | 0.10 | 0.20 | 0.20 | - |
| **Routine hospital data** | | | | | | | |
| Outpatient attendance during study (D) | 28,613 | 59,853 | 5,403 | 1,278 | 48,296 | 36,854 | - |
| Estimated number eligible (C*D = E) | 5,867 | 11,073 | 1,891 | 128 | 9,660 | 7,371 | - |
| **Study data** | | | | | | | |
| Number of outpatients recruited (F) | 175 | 183 | 87 | 29 | 147 | 147 | - |
| Outpatient weighting (1 : E/F) | 1 : 34 | 1 : 64 | 1 : 22 | 1 : 5 | 1 : 66 | 1 : 50 | - |
| **Outcomes** | | | | | | | |
| Frequency | 39/546 | 36/816 | 0/205 | 0/77 | 32/938 | 26/611 | |
| Unweighted prevalence (95% CI) | 7.1% (5.1-9.6) | 4.4% (3.1-6.1) | - | - | 3.4% (2.3-4.8) | 4.3% (2.8-6.2) | **3.9% (3.3-4.7)** |
| Weighted prevalence (95% CI) | 0.64% (0.45-0.90) | 0.30% (0.20-0.42) | - | - | 0.31% (0.21-0.45) | 0.39% (0.26-0.59) | **0.34% (0.28-0.41)** |

Due to high numbers of outpatients, consecutive enrolment of outpatients was not feasible, and recruitment was stratified by admission status, with consecutive screening of inpatients, whilst outpatients were randomly selected for screening. To account for this, the outpatient strata was weighted in the analyses. For one week every 4-6 months, outpatient recruitment was paused and consecutive attendances screened for eligibility. These screening week data were triangulated with routinely collected hospital attendance data to estimate the total number of eligible outpatients presenting to each site during the recruitment period, to determine the weights to be used in the analyses.

Accordingly, screening weeks were planned at each site where outpatient screening was randomised. At study inception outpatient screening was randomised at all sites. Following the start of the Covid-19 pandemic, attendance rates at the site in Indonesia decreased substantially, such that consecutive outpatient screening became possible. The switch to consecutive screening in Indonesia occurred on 10/04/2021 (three weeks after site initiation), covering 90.4% (66/73) of outpatient recruitment at that site. As consecutive screening was used for the majority of outpatient recruitment in Indonesia, no weighting was necessary. However, to align with the methodology used at the other sites, where the estimated number of eligible outpatients presenting to the study site during the recruitment period (E) assumed no refusals, the Indonesian outpatient data were weighted by the observed outpatient refusal rate (62/135; 46%), to provide a weighting of 1:2 for the Indonesian outpatient data.

The second Viet Nam site joined the study on 8 December 2021 in order to boost recruitment, which had been delayed by the Covid-19 pandemic (appendix p10-11). Due to limited remaining resources at this stage of the study, it was not possible to recruit outpatients at this site. Therefore, outpatient data from the first Viet Nam site are used as a proxy. Weighting of these data for the second Viet Nam site was performed using the ratio of inpatients recruited at each Vietnamese site (612:802) and the total number of outpatient attendances at the first Viet Nam site (48,296), to estimate the number of outpatients that would have presented to the second Viet Nam site: 48,296 * (612/802) = 36,854. Assuming the same proportion (0.20) of eligible outpatients at both Viet Nam sites, the number of eligible outpatients presenting to the second Viet Nam site was estimated as: 36,854 * 0.20 = 7,371. This number was then used to determine the weighting to apply to the outpatient data from the first Viet Nam site, as a proxy for outpatient data at the second Viet Nam site: 7,371/147 = 50. Key assumptions underlying this are that the ratio of inpatients to outpatients, proportion of eligible outpatients, and profile of outpatients are similar at both Vietnamese sites.

### 7. Table S6: Candidate predictors entered into the prediction models.

| Parameter (units) | Method / Definition | Handling |
|---|---|---|
| Age (months) | Interview with caregiver | Continuous |
| Mid-upper arm circumference (mm) | *Médecins Sans Frontières* traffic light MUAC tape | Continuous |
| Heart rate (bpm) | Massimo Rad-5v pulse oximeter with paediatric and neonatal probes | Continuous |
| Respiratory rate (bpm) | Manual count for 60 seconds using clicker counter and timer | Continuous |
| Axillary temperature (°C) | Digital thermometer: operating range 32.0-42.9°C; accuracy ± 0.1°C | Continuous |
| Mental state (AVPU scale) | Alert (A) vs. not alert (V or P or U) | Binary |
| Capillary refill time | Pressure applied to sternum for five seconds; prolonged > 2 seconds | Binary |
| Recent hospitalisation | Interview with caregiver; overnight admission within last 6 months | Binary |
| Convulsions | Interview with caregiver; period of unresponsiveness followed by stiffening of the limbs and/or repetitive, rhythmic movements of a part of the body | Binary |
| Prostration | Inability to feed, sit, stand, or walk when previously able | Binary |
| Intractable vomiting | Interview with caregiver; vomited after every feed or drink in last 12h | Binary |
| Oxygen saturation in room air (%) | Massimo Rad-5v pulse oximeter with paediatric and neonatal probes | Continuous |
| ANG-1 (pg/ml) | Simple Plex Ella microfluidic platform (EDTA-plasma) | Continuous |
| ANG-2 (pg/ml) | Simple Plex Ella microfluidic platform (EDTA-plasma) | Continuous |
| sFLT-1 (pg/ml) | Simple Plex Ella microfluidic platform (EDTA-plasma) | Continuous |
| CHI3L1 (ng/ml) | Simple Plex Ella microfluidic platform (EDTA-plasma) | Continuous |
| CRP (mg/l) | Simple Plex Ella microfluidic platform (EDTA-plasma) | Continuous |
| IL-1ra (pg/ml) | Simple Plex Ella microfluidic platform (EDTA-plasma) | Continuous |
| IL-6 (pg/ml) | Simple Plex Ella microfluidic platform (EDTA-plasma) | Continuous |
| IL-8 (pg/ml) | Simple Plex Ella microfluidic platform (EDTA-plasma) | Continuous |
| IL-10 (pg/ml) | Simple Plex Ella microfluidic platform (EDTA-plasma) | Continuous |
| IP-10 (pg/ml) | Simple Plex Ella microfluidic platform (EDTA-plasma) | Continuous |
| PCT (ng/ml) | Simple Plex Ella microfluidic platform (EDTA-plasma) | Continuous |
| sTNF-R1 (pg/ml) | Simple Plex Ella microfluidic platform (EDTA-plasma) | Continuous |
| sTREM1 (pg/ml) | Simple Plex Ella microfluidic platform (EDTA-plasma) | Continuous |
| suPAR (ng/ml) | suPARnostic ELISA (EDTA-plasma) | Continuous |
| Lactate (mmol/l) | LACT2 platform (fluoride-oxalate plasma) | Continuous |
| Glucose (mmol/l) | GLUC3 platform (fluoride-oxalate plasma) | Continuous |
| Hb (g/dL) | Local hospital laboratory platform (EDTA whole blood) | Continuous |

### 8. Figure S1: Cost-effectiveness analysis decision tree model structure

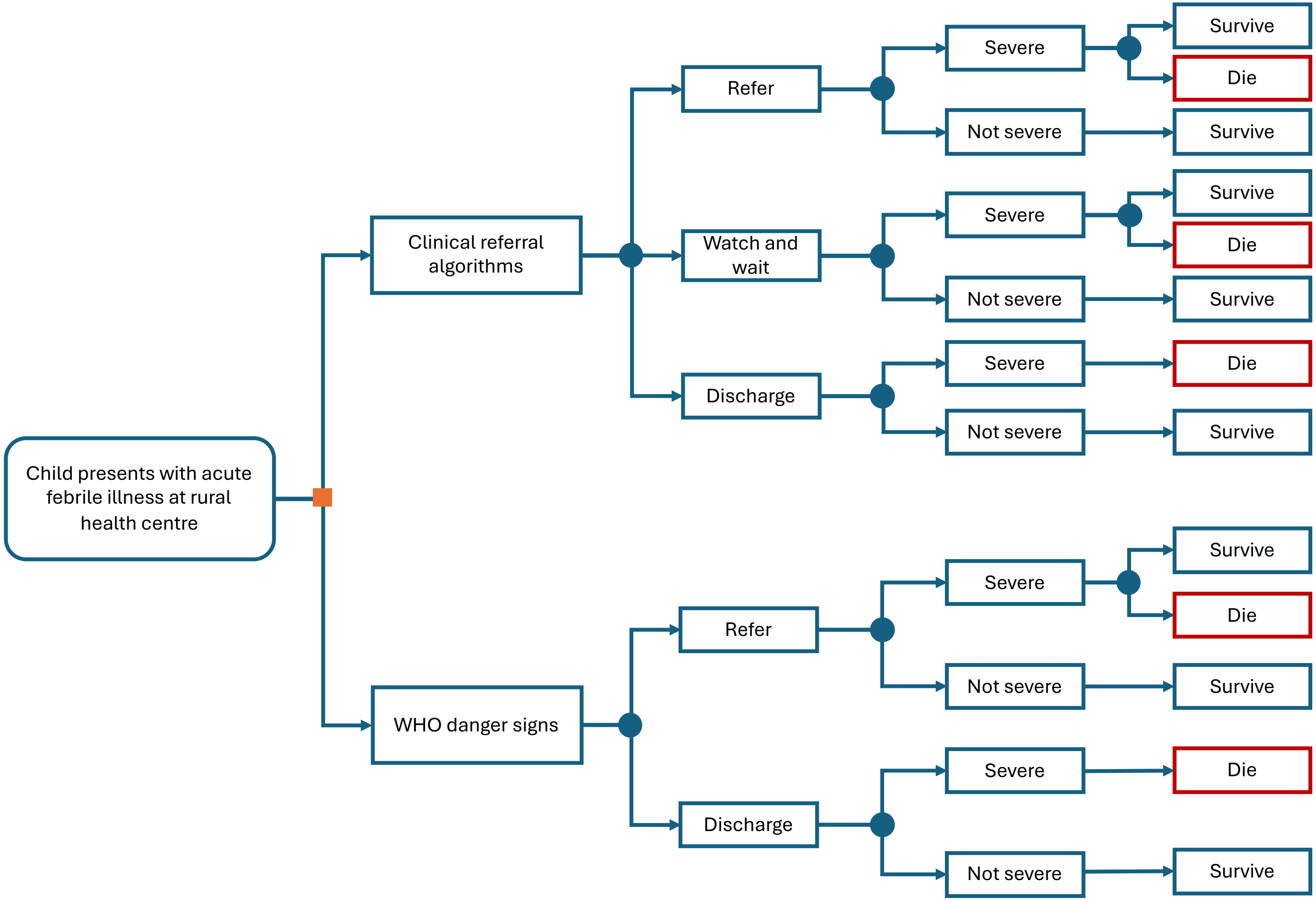

### 9. Table S7: Parameters for cost-effectiveness analyses.

| Parameter | Value | Source | Notes |
|---|---|---|---|
| Average population age | 1 year | Primary data | 16.8 months, rounded to 1 year |
| Life expectancy at population age | 75.21 years | UN Population Prospects 2024[27] | Life expectancy at age x - Bangladesh |
| Percentage of patients that will develop severe symptoms | 0.3% | Primary data | 133/3405 patients (weighted) |
| Mortality if severe and referred | 16.54% | Primary data | 22/133 patients |
| Mortality if severe and not referred | 100% | Assumed | |
| $SpO_2$ cost per patient | $0.45 | Chew et al. 2022[28] | $275 purchase cost and $55 maintenance cost. Assumed two patients per day with a useful lifespan of one year |
| sTREM1 cost per patient | $3.87 | Calarco et al. 2023[29] | Assume cost same as CRP. Used the median cost of analyser ($1000) and consumables ($2.50). Assume used by 2 patients per day with a useful lifespan of one year |
| Outpatient appointment cost per patient for initial assessment | $2.80 | WHO-CHOICE[30] | Outpatient day cost at a primary hospital |
| Outpatient appointment cost per patient for watch and wait | $2.80 | WHO-CHOICE[30] | Outpatient day cost at a primary hospital |
| Inpatient day cost per patient at tertiary hospital | $12.90 | WHO-CHOICE[30] | Inpatient day cost at a tertiary hospital |
| Vital organ support cost per patient (entire duration of stay) | $1,658.90 | Purba et al. 2020[31] | Cost per surviving sepsis patient |
| Referral cost per patient | $12.81 | Nanyonjo et al. 2015[32] | |
| Length of stay for non-severe referred patients | 3 days | Primary data | 2-4 days |

### 10. Table S8: Ethical approvals.

| Ethical Review Board | Approval Reference | Country |
|---|---|---|
| *Médecins Sans Frontières* Ethical Review Board | MSF ERB 1967 | NA |
| Oxford Tropical Medicine Research Committee | OxTREC 59-19 | NA |
| International Centre for Diarrhoeal Disease Research | PR-200006 | Bangladesh |
| Angkor Hospital for Children Research Committee | 01296/19AHC | Cambodia |
| National Ethics Committee for Health Research | 264/NECHR | Cambodia |
| Medical and Health Research Ethics Committee | KE/FK/1397/EC/2019 | Indonesia |
| National Ethics Committee for Health Research | 051/NECHR | Laos |
| University of Medicine and Pharmacy at Ho Chi Minh City | 818/HDDD-DHYD | Viet Nam |
| Ethics Committee for Biomedical Research | VNCH-RICH-2021-77 | Viet Nam |

### 11. Table S9: TRIPOD checklist.

| Section/Topic | | | Checklist Item | Page |
|---|---|---|---|---|
| **Title and abstract** | | | | |
| Title | 1 | D;V | Identify the study as developing and/or validating a multivariable prediction model, the target population, and the outcome to be predicted. | 1 |
| Abstract | 2 | D;V | Provide a summary of objectives, study design, setting, participants, sample size, predictors, outcome, statistical analysis, results, and conclusions. | 3 |
| **Introduction** | | | | |
| Background and objectives | 3a | D;V | Explain the medical context (including whether diagnostic or prognostic) and rationale for developing or validating the multivariable prediction model, including references to existing models. | 4-5 |
| | 3b | D;V | Specify the objectives, including whether the study describes the development or validation of the model or both. | 5 |
| **Methods** | | | | |
| Source of data | 4a | D;V | Describe the study design or source of data (e.g., randomized trial, cohort, or registry data), separately for the development and validation data sets, if applicable. | 15 |
| | 4b | D;V | Specify the key study dates, including start of accrual; end of accrual; and, if applicable, end of follow-up. | 6 |
| Participants | 5a | D;V | Specify key elements of the study setting (e.g., primary care, secondary care, general population) including number and location of centers. | 15 |
| | 5b | D;V | Describe eligibility criteria for participants. | 15 |
| | 5c | D;V | Give details of treatments received, if relevant. | 16 |
| Outcome | 6a | D;V | Clearly define the outcome that is predicted by the prediction model, including how and when assessed. | 18 |
| | 6b | D;V | Report any actions to blind assessment of the outcome to be predicted. | NA |
| Predictors | 7a | D;V | Clearly define all predictors used in developing or validating the multivariable prediction model, including how and when they were measured. | 16; Table S2 |
| | 7b | D;V | Report any actions to blind assessment of predictors for the outcome and other predictors. | NA |
| Sample size | 8 | D;V | Explain how the study size was arrived at. | 18 |
| Missing data | 9 | D;V | Describe how missing data were handled (e.g., complete-case analysis, single imputation, multiple imputation) with details of any imputation method. | 19 |
| Statistical analysis methods | 10a | D | Describe how predictors were handled in the analyses. | 19 |
| | 10b | D | Specify type of model, all model-building procedures (including any predictor selection), and method for internal validation. | 19 |
| | 10c | V | For validation, describe how the predictions were calculated. | 20 |
| | 10d | D;V | Specify all measures used to assess model performance and, if relevant, to compare multiple models. | 20 |
| | 10e | V | Describe any model updating (e.g., recalibration) arising from the validation, if done. | NA |
| Risk groups | 11 | D;V | Provide details on how risk groups were created, if done. | 20 |
| Development vs. validation | 12 | V | For validation, identify any differences from the development data in setting, eligibility criteria, outcome, and predictors. | 7-8 |
| **Results** | | | | |
| Participants | 13a | D;V | Describe the flow of participants through the study, including the number of participants with and without the outcome and, if applicable, a summary of the follow-up time. A diagram may be helpful. | 6; Fig S1 |
| | 13b | D;V | Describe the characteristics of the participants (basic demographics, clinical features, available predictors), including the number of participants with missing data for predictors and outcome. | 6-7; Table 1 |
| | 13c | V | For validation, show a comparison with the development data of the distribution of important variables (demographics, predictors and outcome). | 6-7; Table 1 |
| Model development | 14a | D | Specify the number of participants and outcome events in each analysis. | 6-7 |
| | 14b | D | If done, report the unadjusted association between each candidate predictor and outcome. | 6-7 |
| Model specification | 15a | D | Present the full prediction model to allow predictions for individuals (i.e., all regression coefficients, and model intercept or baseline survival at a given time point). | Table S6 |
| | 15b | D | Explain how to the use the prediction model. | Table S6 |
| Model performance | 16 | D;V | Report performance measures (with CIs) for the prediction model. | 7-9; Table 2 |
| Model-updating | 17 | V | If done, report the results from any model updating (i.e., model specification, model performance). | NA |
| **Discussion** | | | | |
| Limitations | 18 | D;V | Discuss any limitations of the study (such as nonrepresentative sample, few events per predictor, missing data). | 12-14 |
| Interpretation | 19a | V | For validation, discuss the results with reference to performance in the development data, and any other validation data. | NA |
| | 19b | D;V | Give an overall interpretation of the results, considering objectives, limitations, results from similar studies, and other relevant evidence. | 11-12; 14 |
| Implications | 20 | D;V | Discuss the potential clinical use of the model and implications for future research. | 14 |
| **Other information** | | | | |
| Supplementary information | 21 | D;V | Provide information about the availability of supplementary resources, such as study protocol, Web calculator, and data sets. | 22 |
| Funding | 22 | D;V | Give the source of funding and the role of the funders for the present study. | 21 |

### 12. Figure S2: Study flowchart.

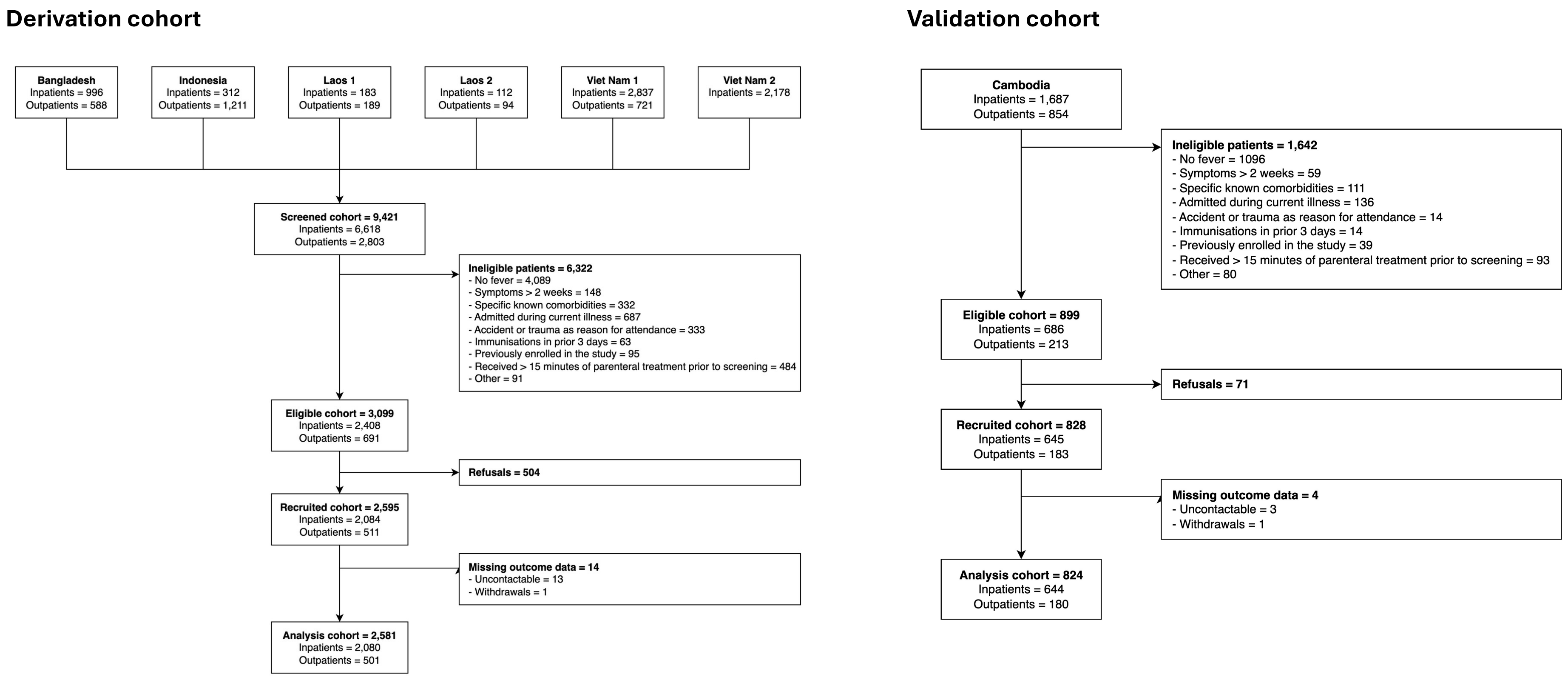

One reason for ineligibility is provided per patient, according to the hierarchy listed in the figures. 1,245 ineligible children had more than one reason for ineligibility (1,245/7,964; 15.6%).

### 13. Table S10: Microbiological causes of infection in the derivation and validation cohorts.

| Characteristic | Overall<br>N = 3,405[1] | Derivation cohort<br>N = 2,581[1] | Validation cohort<br>N = 824[1] |
|---|---|---|---|
| Zika | 8 / 3,289 (0.2%) | 1 / 2,474 (<0.1%) | 7 / 815 (0.9%) |
| Dengue | 109 / 3,289 (3.3%) | 107 / 2,474 (4.3%) | 2 / 815 (0.2%) |
| Chikungunya | 47 / 3,289 (1.4%) | 4 / 2,474 (0.2%) | 43 / 815 (5.3%) |
| *Leptospirosis* spp. | 4 / 3,128 (0.1%) | 4 / 2,313 (0.2%) | 0 / 815 (0%) |
| *Rickettsia* spp. | 6 / 3,128 (0.2%) | 5 / 2,313 (0.2%) | 1 / 815 (0.1%) |
| *Orientia tsutsugamushi* | 3 / 3,128 (<0.1%) | 1 / 2,313 (<0.1%) | 2 / 815 (0.2%) |
| SARS-CoV-2 | 81 / 2,861 (2.8%) | 74 / 2,209 (3.3%) | 7 / 652 (1.1%) |
| Influenza A | 87 / 2,861 (3.0%) | 69 / 2,209 (3.1%) | 18 / 652 (2.8%) |
| Influenza B | 59 / 2,861 (2.1%) | 59 / 2,209 (2.7%) | 0 / 652 (0%) |
| Respiratory syncytial virus | 429 / 2,861 (15%) | 349 / 2,209 (16%) | 80 / 652 (12%) |
| Human metapneumovirus | 65 / 2,209 (2.9%) | 65 / 2,209 (2.9%) | 0 / 0 |
| *Mycoplasma pneumoniae* | 3 / 2,209 (0.1%) | 3 / 2,209 (0.1%) | 0 / 0 |
| *Chlamydia pneumoniae* | 3 / 2,209 (0.1%) | 3 / 2,209 (0.1%) | 0 / 0 |
| *Bordetella parapertussis* | 8 / 2,209 (0.4%) | 8 / 2,209 (0.4%) | 0 / 0 |
| *Bordetella pertussis* | 1 / 2,209 (<0.1%) | 1 / 2,209 (<0.1%) | 0 / 0 |
| Bacteraemia* | 19 / 1,192 (1.6%) | 12 / 781 (1.5%) | 7 / 411 (1.7%) |

[1]n / N (%)

*Bacteraemia diagnosed by blood culture: *Staphylococcus aureus*, n = 5; *Escherichia coli*, n = 3; *Acinetobacter* spp., n = 2; *Salmonella* spp., n = 2; Other, n = 7 (*Campylobacter jejuni*, n = 1; *Enterobacter* spp., n = 1; *Enterococcus faecalis*, n = 1; *Haemophilus influenzae*, n = 1; *Streptococcus pneumoniae*, n = 1; *Streptococcus pyogenes*, n = 1; *Vibrio* spp., n = 1). In Cambodia (validation cohort), respiratory samples were tested for SARS-CoV-2, influenza A, influenza B, and RSV. In Indonesia (part of the derivation cohort), PCR testing was performed for Zika, dengue, and chikungunya. When feasible, the study supported collection of blood cultures at the discretion of the treating clinical team.

14. **Table S11: Variable selection and performance of the *clinical-biomarker models* when each biomarker was included in turn alongside the candidate clinical predictors.**

| | sTREM1 | CRP | PCT | ANG-2 | sTNF-R1 | sFLT-1 | Hb | IL-6 | IL-1ra | CHI3L1 | suPAR | IL-10 | IL-8 | IP-10 | ANG-1 | Glucose | Lactate |
|---|---|---|---|---|---|---|---|---|---|---|---|---|---|---|---|---|---|
| **Age (months)** | red | red | red | red | red | red | red | red | red | red | red | red | red | red | red | red | red |
| **MUAC (mm)** | red | red | red | red | red | red | red | red | red | red | red | red | red | red | red | red | red |
| **Heart rate (bpm)** | green | green | green | green | green | green | green | green | green | green | green | green | green | green | green | green | green |
| **Respiratory rate (bpm)** | green | green | green | green | green | green | green | green | green | green | green | green | green | green | green | green | green |
| **Temperature (°C)** | red | green | red | red | red | red | red | red | red | red | red | red | red | red | red | red | red |
| **Altered mental state** | green | green | green | green | green | green | green | green | green | green | green | green | green | green | green | green | green |
| **Prolonged CRT** | red | green | green | green | green | green | green | green | green | green | green | green | green | green | green | green | green |
| **Recent hospitalisation** | red | red | red | red | red | red | red | red | red | red | red | red | red | red | red | red | red |
| **Prostration** | green | green | green | green | green | green | green | green | green | green | green | green | green | green | green | green | green |
| **Intractable vomiting** | green | green | green | green | green | green | green | green | green | green | green | green | green | green | green | green | green |
| **Convulsions** | red | red | red | red | red | red | red | red | red | red | red | red | red | red | red | red | red |
| **sTREM1 (pg/ml)** | green | yellow | yellow | yellow | yellow | yellow | yellow | yellow | yellow | yellow | yellow | yellow | yellow | yellow | yellow | yellow | yellow |
| **CRP (mg/l)** | yellow | green | yellow | yellow | yellow | yellow | yellow | yellow | yellow | yellow | yellow | yellow | yellow | yellow | yellow | yellow | yellow |
| **PCT (ng/ml)** | yellow | yellow | green | yellow | yellow | yellow | yellow | yellow | yellow | yellow | yellow | yellow | yellow | yellow | yellow | yellow | yellow |
| **ANG-2 (pg/ml)** | yellow | yellow | yellow | green | yellow | yellow | yellow | yellow | yellow | yellow | yellow | yellow | yellow | yellow | yellow | yellow | yellow |
| **sTNF-R1 (pg/ml)** | yellow | yellow | yellow | yellow | green | yellow | yellow | yellow | yellow | yellow | yellow | yellow | yellow | yellow | yellow | yellow | yellow |
| **sFLT-1 (pg/ml)** | yellow | yellow | yellow | yellow | yellow | green | yellow | yellow | yellow | yellow | yellow | yellow | yellow | yellow | yellow | yellow | yellow |
| **Hb (g/dl)** | yellow | yellow | yellow | yellow | yellow | yellow | green | yellow | yellow | yellow | yellow | yellow | yellow | yellow | yellow | yellow | yellow |
| **IL-6 (pg/ml)** | yellow | yellow | yellow | yellow | yellow | yellow | yellow | green | yellow | yellow | yellow | yellow | yellow | yellow | yellow | yellow | yellow |
| **IL-1ra (pg/ml)** | yellow | yellow | yellow | yellow | yellow | yellow | yellow | yellow | green | yellow | yellow | yellow | yellow | yellow | yellow | yellow | yellow |
| **CHI3L1 (ng/ml)** | yellow | yellow | yellow | yellow | yellow | yellow | yellow | yellow | yellow | green | yellow | yellow | yellow | yellow | yellow | yellow | yellow |
| **suPAR (ng/ml)** | yellow | yellow | yellow | yellow | yellow | yellow | yellow | yellow | yellow | yellow | green | yellow | yellow | yellow | yellow | yellow | yellow |
| **IL-10 (pg/ml)** | yellow | yellow | yellow | yellow | yellow | yellow | yellow | yellow | yellow | yellow | yellow | red | yellow | yellow | yellow | yellow | yellow |
| **IL-8 (pg/ml)** | yellow | yellow | yellow | yellow | yellow | yellow | yellow | yellow | yellow | yellow | yellow | yellow | red | yellow | yellow | yellow | yellow |
| **IP-10 (pg/ml)** | yellow | yellow | yellow | yellow | yellow | yellow | yellow | yellow | yellow | yellow | yellow | yellow | yellow | red | yellow | yellow | yellow |
| **ANG-1 (pg/ml)** | yellow | yellow | yellow | yellow | yellow | yellow | yellow | yellow | yellow | yellow | yellow | yellow | yellow | yellow | red | yellow | yellow |
| **Glucose (mmol/l)** | yellow | yellow | yellow | yellow | yellow | yellow | yellow | yellow | yellow | yellow | yellow | yellow | yellow | yellow | yellow | red | yellow |
| **Lactate (mmol/l)** | yellow | yellow | yellow | yellow | yellow | yellow | yellow | yellow | yellow | yellow | yellow | yellow | yellow | yellow | yellow | yellow | red |
| **AUC (optimism adjusted)** | 0.93 | 0.93 | 0.91 | 0.92 | 0.92 | 0.91 | 0.91 | 0.91 | 0.90 | 0.90 | 0.91 | 0.91 | 0.91 | 0.91 | 0.91 | 0.91 | 0.91 |

Green indicates variable was selected, red indicates variable was eliminated, yellow indicates variable was not entered into the model. Model performance is quantified in the derivation cohort using the optimism adjusted weighted AUC

**15. Table S12: Variable selection and performance of the *clinical-biomarker models* when each biomarker was included in turn alongside the candidate clinical predictors and $SpO_2$.**

| | sTREM1 | CRP | PCT | ANG-2 | sTNF-R1 | sFLT-1 | Hb | IL-6 | IL-1ra | CHI3L1 | suPAR | IL-10 | IL-8 | IP-10 | ANG-1 | Glucose | Lactate |
|---|---|---|---|---|---|---|---|---|---|---|---|---|---|---|---|---|---|
| **Age (months)** | | | | | | | | | | | | | | | | | |
| **MUAC (mm)** | | | | | | | | | | | | | | | | | |
| **Heart rate (bpm)** | | | | | | | | | | | | | | | | | |
| **Respiratory rate (bpm)** | | | | | | | | | | | | | | | | | |
| **Temperature (°C)** | | | | | | | | | | | | | | | | | |
| **Altered mental state** | | | | | | | | | | | | | | | | | |
| **Prolonged CRT** | | | | | | | | | | | | | | | | | |
| **Recent hospitalisation** | | | | | | | | | | | | | | | | | |
| **Prostration** | | | | | | | | | | | | | | | | | |
| **Intractable vomiting** | | | | | | | | | | | | | | | | | |
| **Convulsions** | | | | | | | | | | | | | | | | | |
| **$SpO_2$ (%)** | | | | | | | | | | | | | | | | | |
| **sTREM1 (pg/ml)** | | | | | | | | | | | | | | | | | |
| **CRP (mg/l)** | | | | | | | | | | | | | | | | | |
| **PCT (ng/ml)** | | | | | | | | | | | | | | | | | |
| **ANG-2 (pg/ml)** | | | | | | | | | | | | | | | | | |
| **sTNF-R1 (pg/ml)** | | | | | | | | | | | | | | | | | |
| **sFLT-1 (pg/ml)** | | | | | | | | | | | | | | | | | |
| **Hb (g/dl)** | | | | | | | | | | | | | | | | | |
| **IL-6 (pg/ml)** | | | | | | | | | | | | | | | | | |
| **IL-1ra (pg/ml)** | | | | | | | | | | | | | | | | | |
| **CHI3L1 (ng/ml)** | | | | | | | | | | | | | | | | | |
| **suPAR (ng/ml)** | | | | | | | | | | | | | | | | | |
| **IL-10 (pg/ml)** | | | | | | | | | | | | | | | | | |
| **IL-8 (pg/ml)** | | | | | | | | | | | | | | | | | |
| **IP-10 (pg/ml)** | | | | | | | | | | | | | | | | | |
| **ANG-1 (pg/ml)** | | | | | | | | | | | | | | | | | |
| **Glucose (mmol/l)** | | | | | | | | | | | | | | | | | |
| **Lactate (mmol/l)** | | | | | | | | | | | | | | | | | |
| **AUC (optimism adjusted)** | 0.94 | 0.94 | 0.93 | 0.92 | 0.92 | 0.92 | 0.92 | 0.92 | 0.92 | 0.92 | 0.91 | 0.91 | 0.91 | 0.91 | 0.91 | 0.91 | 0.91 |

Green indicates variable was selected, red indicates variable was eliminated, yellow indicates variable was not entered into the model. Model performance is quantified in the derivation cohort using the optimism adjusted weighted AUC.

16. **Table S13: Variable selection and performance of the *clinical-biomarker models* when each biomarker was included in turn alongside the candidate clinical predictors, excluding the northern Viet Nam site.**

| | sTREM1 | CRP | PCT | ANG-2 | sTNF-R1 | sFLT-1 | Hb | IL-6 | IL-1ra | CHI3L1 | suPAR | IL-10 | IL-8 | IP-10 | ANG-1 | Glucose | Lactate |
|---|---|---|---|---|---|---|---|---|---|---|---|---|---|---|---|---|---|
| **Age (months)** | red | red | red | red | red | red | red | red | red | red | red | red | red | red | red | red | red |
| **MUAC (mm)** | red | red | red | red | red | red | red | red | red | red | red | red | red | red | red | red | red |
| **Heart rate (bpm)** | green | green | green | green | green | green | green | green | green | green | green | green | green | green | green | green | green |
| **Respiratory rate (bpm)** | green | green | green | green | green | green | green | green | green | green | green | green | green | green | green | green | green |
| **Temperature (°C)** | red | red | red | red | red | red | red | red | red | red | red | red | red | red | red | red | red |
| **Altered mental state** | red | red | red | red | red | red | red | red | red | red | red | red | red | red | red | red | red |
| **Prolonged CRT** | red | red | red | red | red | red | red | red | red | red | red | red | red | red | red | red | red |
| **Recent hospitalisation** | red | red | red | red | red | red | red | red | red | red | red | red | red | red | red | red | red |
| **Prostration** | green | green | green | green | green | green | green | green | green | green | green | green | green | green | green | green | green |
| **Intractable vomiting** | green | green | green | green | green | green | green | green | green | green | green | green | green | green | green | green | green |
| **Convulsions** | green | green | green | green | green | red | green | green | green | green | green | green | green | green | green | green | green |
| **$SpO_2$ (%)** | green | green | green | green | green | green | green | green | green | green | green | green | green | green | green | green | green |
| **sTREM1 (pg/ml)** | green | yellow | yellow | yellow | yellow | yellow | yellow | yellow | yellow | yellow | yellow | yellow | yellow | yellow | yellow | yellow | yellow |
| **CRP (mg/l)** | yellow | red | yellow | yellow | yellow | yellow | yellow | yellow | yellow | yellow | yellow | yellow | yellow | yellow | yellow | yellow | yellow |
| **PCT (ng/ml)** | yellow | yellow | green | yellow | yellow | yellow | yellow | yellow | yellow | yellow | yellow | yellow | yellow | yellow | yellow | yellow | yellow |
| **ANG-2 (pg/ml)** | yellow | yellow | yellow | green | yellow | yellow | yellow | yellow | yellow | yellow | yellow | yellow | yellow | yellow | yellow | yellow | yellow |
| **sTNF-R1 (pg/ml)** | yellow | yellow | yellow | yellow | green | yellow | yellow | yellow | yellow | yellow | yellow | yellow | yellow | yellow | yellow | yellow | yellow |
| **sFLT-1 (pg/ml)** | yellow | yellow | yellow | yellow | yellow | green | yellow | yellow | yellow | yellow | yellow | yellow | yellow | yellow | yellow | yellow | yellow |
| **Hb (g/dl)** | yellow | yellow | yellow | yellow | yellow | yellow | green | yellow | yellow | yellow | yellow | yellow | yellow | yellow | yellow | yellow | yellow |
| **IL-6 (pg/ml)** | yellow | yellow | yellow | yellow | yellow | yellow | yellow | red | yellow | yellow | yellow | yellow | yellow | yellow | yellow | yellow | yellow |
| **IL-1ra (pg/ml)** | yellow | yellow | yellow | yellow | yellow | yellow | yellow | yellow | red | yellow | yellow | yellow | yellow | yellow | yellow | yellow | yellow |
| **CHI3L1 (ng/ml)** | yellow | yellow | yellow | yellow | yellow | yellow | yellow | yellow | yellow | red | yellow | yellow | yellow | yellow | yellow | yellow | yellow |
| **suPAR (ng/ml)** | yellow | yellow | yellow | yellow | yellow | yellow | yellow | yellow | yellow | yellow | green | yellow | yellow | yellow | yellow | yellow | yellow |
| **IL-10 (pg/ml)** | yellow | yellow | yellow | yellow | yellow | yellow | yellow | yellow | yellow | yellow | yellow | red | yellow | yellow | yellow | yellow | yellow |
| **IL-8 (pg/ml)** | yellow | yellow | yellow | yellow | yellow | yellow | yellow | yellow | yellow | yellow | yellow | yellow | red | yellow | yellow | yellow | yellow |
| **IP-10 (pg/ml)** | yellow | yellow | yellow | yellow | yellow | yellow | yellow | yellow | yellow | yellow | yellow | yellow | yellow | red | yellow | yellow | yellow |
| **ANG-1 (pg/ml)** | yellow | yellow | yellow | yellow | yellow | yellow | yellow | yellow | yellow | yellow | yellow | yellow | yellow | yellow | red | yellow | yellow |
| **Glucose (mmol/l)** | yellow | yellow | yellow | yellow | yellow | yellow | yellow | yellow | yellow | yellow | yellow | yellow | yellow | yellow | yellow | red | yellow |
| **Lactate (mmol/l)** | yellow | yellow | yellow | yellow | yellow | yellow | yellow | yellow | yellow | yellow | yellow | yellow | yellow | yellow | yellow | yellow | red |
| **AUC (optimism adjusted)** | 0.98 | 0.97 | 0.97 | 0.98 | 0.97 | 0.97 | 0.98 | 0.97 | 0.97 | 0.97 | 0.97 | 0.97 | 0.97 | 0.97 | 0.97 | 0.97 | 0.97 |

Green indicates variable was selected, red indicates variable was eliminated, yellow indicates variable was not entered into the model. Model performance is quantified in the derivation cohort using the optimism adjusted weighted AUC.

17. **Tables S14: Logistic regression equations for the four clinical prediction models.**

**Instructions to use these equations:**

1. Input the value for each variable.
2. Compute the linear predictor (LP), which is the value of the right-hand side of the equation.
3. Convert the LP to a probability using the logistic function: p = 1 / (1 + e^(-LP))
4. The result p is the predicted probability

**Clinical model**

| **Variable** | **Coefficient (β)** | **Odds Ratio (95% CI)** |
|---|---|---|
| Constant | -13.8215 | - |
| Intractable vomiting | 1.1946 | 3.30 (1.78-6.13) |
| Prostration | 2.1689 | 8.75 (4.26-17.97) |
| Heart rate | 0.0322 | 1.03 (1.02-1.05) |
| Respiratory rate | 0.0624 | 1.06 (1.04-1.09) |
| Prolonged capillary refill time | 1.0570 | 2.88 (1.26-6.58) |
| Altered mental state | 1.9074 | 6.74 (2.06-22.07) |

**Logistic Regression Equation:** log(p / (1 - p)) = -13.8215 + (1.1946*vomit) + (2.1689*prostration) + (0.0322*heart_rate) + (0.0624*respiratory_rate) + (1.0570*cap_refill_time) + (1.9074*mental_state)

**Pulse oximetry model**

| **Variable** | **Coefficient (β)** | **Odds Ratio (95% CI)** |
|---|---|---|
| Constant | 1.9561 | - |
| Convulsions | 1.0210 | 2.78 (1.23-6.27) |
| Intractable vomiting | 1.2767 | 3.58 (2.02-6.37) |
| Prostration | 2.1623 | 8.69 (4.45-16.97) |
| Heart rate | 0.0268 | 1.03 (1.01-1.04) |
| Respiratory rate | 0.0624 | 1.06 (1.04-1.09) |
| $SpO_2$ | -0.1541 | 0.86 (0.80-0.92) |

**Logistic Regression Equation:** log(p / (1 - p)) = 1.9561 + (1.0210*convulsion) + (1.2767*vomit) + (2.1623*prostration) + (0.0268*heart_rate) + (0.0624*respiratory_rate) – (0.1541*$SpO_2$)

**sTREM1 model**

| **Variable** | **Coefficient (β)** | **Odds Ratio (95% CI)** |
|---|---|---|
| Constant | -14.5167 | - |
| Intractable vomiting | 1.2290 | 3.42 (1.90-6.16) |
| Prostration | 1.9687 | 7.16 (3.66-14.02) |
| Heart rate | 0.0342 | 1.03 (1.02-1.05) |
| Respiratory rate | 0.0540 | 1.06 (1.03-1.09) |
| Altered mental state | 2.0153 | 7.50 (2.30-24.43) |
| sTREM1 | 0.0030 | 1.00 (1.00-1.00) |

**Logistic Regression Equation:** log(p / (1 - p)) = -14.5167 + (1.2290*vomit) + (1.9687*prostration) + (0.0342*heart_rate) + (0.0540*respiratory_rate) + (2.0153*mental_state) + (0.0030*sTREM1)

**Combined model**

| Variable | Coefficient (β) | Odds Ratio (95% CI) |
|---|---|---|
| Constant | 0.5857 | - |
| Prostration | 1.7803 | 5.93 (2.90-12.14) |
| Intractable vomiting | 1.3179 | 3.74 (2.07-6.74) |
| Heart rate | 0.0290 | 1.03 (1.02-1.04) |
| Respiratory rate | 0.0504 | 1.05 (1.02-1.08) |
| sTREM1 | 0.0029 | 1.00 (1.00-1.00) |
| $SpO_2$ | -0.1454 | 0.86 (0.80-0.93) |

**Logistic Regression Equation:** log(p / (1 - p)) = 0.5857 + (1.7803*prostration) + (1.3179*vomit) + (0.0290*heart_rate) + (0.0504*respiratory_rate) + (0.0029*sTREM1) – (0.1454*$SpO_2$)

### 18. Table S15: Characteristics of participants who developed severe disease, stratified by whether they were identified by each prediction model (derivation and validation cohorts pooled).

| Characteristic | Severe patients N = 133[1] | Clinical model | | Pulse oximetry model | | sTREM1 model | | Combined model | |
|---|---|---|---|---|---|---|---|---|---|
| | | Identified N = 102[1] | Missed N = 31[1] | Identified N = 107[1] | Missed N = 26[1] | Identified N = 111[1] | Missed N = 22[1] | Identified N = 110[1] | Missed N = 23[1] |
| **Demographics and background** | | | | | | | | | |
| Age (months) | 4.9 (2.6, 17.3) | 4.5 (2.5, 11.2) | 25.3 (4.9, 43.9) | 4.5 (2.1, 12.3) | 30.2 (6.3, 48.1) | 4.5 (2.1, 12.7) | 29.0 (6.3, 49.7) | 4.5 (2.5, 12.7) | 27.7 (4.9, 49.7) |
| Male sex | 45 (34%) | 30 (29%) | 15 (48%) | 31 (29%) | 14 (54%) | 34 (31%) | 11 (50%) | 33 (30%) | 12 (52%) |
| **Anthropometrics** | | | | | | | | | |
| Wasted (WHZ < -2)* | 34 (26%) | 30 (29%) | 4 (13%) | 29 (27%) | 5 (19%) | 30 (27%) | 4 (18%) | 30 (28%) | 4 (17%) |
| Stunted (HAZ < -2) | 35 (26%) | 27 (26%) | 8 (26%) | 32 (30%) | 3 (12%) | 32 (29%) | 3 (14%) | 32 (29%) | 3 (13%) |
| MUAC-for-age z-score[a] | -0.5 (-1.5, 0.6) | -0.9 (-1.7, 0.3) | -0.2 (-0.8, 0.9) | -0.7 (-1.6, 0.4) | -0.2 (-0.9, 0.9) | -0.7 (-1.6, 0.4) | 0.0 (-0.8, 0.9) | -0.7 (-1.6, 0.5) | -0.2 (-0.8, 0.9) |
| **Illness characteristics** | | | | | | | | | |
| Duration of illness (days) | 3.0 (2.0, 5.0) | 4.0 (3.0, 5.0) | 2.0 (1.0, 3.0) | 4.0 (3.0, 5.0) | 2.0 (1.0, 4.0) | 4.0 (3.0, 5.0) | 2.0 (1.0, 3.0) | 4.0 (3.0, 5.0) | 2.0 (1.0, 4.0) |
| URTI | 39 (29%) | 29 (28%) | 10 (32%) | 33 (31%) | 6 (23%) | 33 (30%) | 6 (27%) | 34 (31%) | 5 (22%) |
| LRTI | 86 (65%) | 76 (75%) | 10 (32%) | 80 (75%) | 6 (23%) | 80 (72%) | 6 (27%) | 81 (74%) | 5 (22%) |
| Diarrhoeal | 15 (11%) | 12 (12%) | 3 (9.7%) | 12 (11%) | 3 (12%) | 12 (11%) | 3 (14%) | 12 (11%) | 3 (13%) |
| Neurological | 14 (11%) | 12 (12%) | 2 (6.5%) | 12 (11%) | 2 (7.7%) | 12 (11%) | 2 (9.1%) | 12 (11%) | 2 (8.7%) |
| No focus | 8 (6.0%) | 7 (6.9%) | 1 (3.2%) | 8 (7.5%) | 0 (0%) | 8 (7.2%) | 0 (0%) | 8 (7.3%) | 0 (0%) |
| **WHO danger signs** | | | | | | | | | |
| Any WHO danger sign* | 95 (72%) | 76 (75%) | 19 (61%) | 80 (75%) | 15 (58%) | 82 (75%) | 13 (59%) | 81 (74%) | 14 (61%) |
| *Prostration* | 48 (36%) | 47 (46%) | 1 (3.2%) | 47 (44%) | 1 (3.8%) | 48 (43%) | 0 (0%) | 47 (43%) | 1 (4.3%) |
| *Intractable vomiting* | 42 (32%) | 32 (31%) | 10 (32%) | 33 (31%) | 9 (35%) | 33 (30%) | 9 (41%) | 33 (30%) | 9 (39%) |
| *Convulsions** | 14 (11%) | 12 (12%) | 2 (6.5%) | 13 (12%) | 1 (3.8%) | 12 (11%) | 2 (9.1%) | 13 (12%) | 1 (4.3%) |
| *Lethargy** | 61 (46%) | 52 (51%) | 9 (29%) | 55 (52%) | 6 (23%) | 58 (53%) | 3 (14%) | 56 (51%) | 5 (22%) |
| **Vital signs** | | | | | | | | | |
| Heart rate (bpm) | | | | | | | | | |
| *1 to 12 months (bpm)* | 171.5 (157.0, 186.0) | 172.0 (160.0, 189.0) | 150.0 (146.0, 175.0) | 172.0 (158.5, 189.5) | 155.0 (143.0, 172.0) | 172.0 (159.0, 189.0) | 148.0 (140.0, 160.0) | 172.0 (158.5, 189.5) | 155.0 (143.0, 172.0) |
| *12 to 60 months (bpm)* | 160.0 (140.0, 177.0) | 172.0 (160.0, 192.0) | 144.0 (130.5, 154.5) | 171.0 (156.0, 190.0) | 141.0 (129.0, 158.0) | 170.0 (158.0, 190.0) | 141.0 (130.5, 150.0) | 170.5 (158.0, 190.0) | 136.0 (127.0, 150.0) |
| Respiratory rate (bpm) | | | | | | | | | |
| *1 to 12 months (bpm)* | 59.5 (48.0, 66.0) | 60.0 (50.0, 68.0) | 42.0 (35.0, 50.0) | 60.0 (50.0, 67.5) | 41.0 (37.5, 44.0) | 60.0 (50.0, 67.0) | 40.0 (35.0, 42.0) | 60.0 (50.0, 67.5) | 41.0 (37.5, 44.0) |
| *1 to 12 months (bpm)* | 39.0 (32.0, 55.0) | 55.0 (40.0, 67.0) | 32.0 (28.5, 37.5) | 54.0 (39.0, 67.0) | 32.0 (28.0, 38.0) | 50.0 (39.0, 65.0) | 31.5 (28.0, 37.5) | 50.0 (39.0, 65.0) | 31.0 (28.0, 37.0) |
| Oxygen saturation (%)* | 97.0 (95.0, 98.0) | 96.0 (93.0, 98.0) | 98.0 (97.0, 98.0) | 96.0 (92.0, 98.0) | 98.0 (97.0, 99.0) | 96.0 (93.0, 98.0) | 98.0 (97.0, 98.0) | 96.0 (93.0, 98.0) | 98.0 (97.0, 99.0) |
| Axillary temperature (°C) | 37.6 (36.9, 38.3) | 37.6 (37.0, 38.3) | 37.5 (36.7, 38.4) | 37.6 (36.9, 38.3) | 37.7 (37.0, 38.4) | 37.6 (36.9, 38.3) | 37.5 (36.8, 38.3) | 37.6 (36.9, 38.3) | 37.5 (36.8, 38.3) |
| CRT > 2 seconds | 29 (22%) | 28 (27%) | 1 (3.2%) | 28 (26%) | 1 (3.8%) | 28 (25%) | 1 (4.5%) | 28 (25%) | 1 (4.3%) |
| Not alert[b] | 19 (14%) | 19 (19%) | 0 (0%) | 18 (17%) | 1 (3.8%) | 19 (17%) | 0 (0%) | 18 (16%) | 1 (4.3%) |
| **Endothelial activation markers** | | | | | | | | | |
| ANG-1 (pg/ml)* | 5,746.0 (3,426.0, 10,543.0) | 6,345.5 (3,658.0, 11,662.0) | 4,562.0 (3,155.0, 6,225.0) | 6,043.0 (3,610.0, 10,369.0) | 5,178.5 (3,155.0, 11,270.0) | 6,049.0 (3,698.0, 10,894.0) | 3,760.5 (2,977.0, 6,225.0) | 5,998.5 (3,614.0, 10,456.0) | 4,899.0 (2,977.0, 11,270.0) |
| ANG-2 (pg/ml)* | 2,532.0 (1,435.0, 4,361.0) | 3,262.0 (1,763.5, 5,185.0) | 1,546.0 (1,044.0, 2,776.0) | 3,249.0 (1,784.0, 5,168.0) | 1,514.0 (1,011.0, 2,272.0) | 3,275.0 (1,784.0, 5,168.0) | 1,410.5 (982.0, 1,905.0) | 3,244.5 (1,763.5, 5,148.5) | 1,494.0 (982.0, 2,272.0) |
| sFLT-1 (pg/ml)* | 258.0 (202.0, 371.0) | 274.0 (222.0, 462.0) | 211.0 (170.0, 273.0) | 274.0 (218.0, 459.0) | 210.5 (165.0, 268.0) | 274.0 (218.0, 459.0) | 208.0 (165.0, 242.0) | 268.5 (212.5, 452.0) | 211.0 (165.0, 273.0) |
| **Immune activation markers** | | | | | | | | | |

| Characteristic | Severe patients N = 133[1] | Clinical model | | Pulse oximetry model | | sTREM1 model | | Combined model | |
|---|---|---|---|---|---|---|---|---|---|
| | | Identified N = 102[1] | Missed N = 31[1] | Identified N = 107[1] | Missed N = 26[1] | Identified N = 111[1] | Missed N = 22[1] | Identified N = 110[1] | Missed N = 23[1] |
| CHI3L1 (ng/ml)* | 37.8 (24.0, 66.4) | 38.9 (25.4, 80.1) | 29.7 (22.4, 49.5) | 38.9 (24.7, 78.1) | 29.9 (23.6, 49.0) | 38.9 (25.5, 75.3) | 29.2 (15.6, 48.5) | 38.9 (24.0, 76.2) | 29.7 (21.4, 49.0) |
| CRP (mg/l)* | 25.8 (5.0, 105.3) | 22.7 (3.1, 80.5) | 42.9 (9.3, 166.8) | 21.8 (3.6, 78.6) | 89.0 (16.2, 200.7) | 23.5 (3.2, 80.0) | 57.8 (9.8, 166.8) | 23.6 (3.8, 80.5) | 42.9 (9.3, 166.8) |
| IL-1ra (pg/ml)* | 2,829.0 (1,211.0, 9,799.0) | 3,509.5 (1,372.0, 11,423.0) | 1,756.0 (779.0, 7,223.0) | 3,247.0 (1,414.0, 10,537.0) | 1,701.0 (904.0, 7,223.0) | 3,228.0 (1,235.0, 10,537.0) | 1,803.0 (904.0, 7,442.0) | 3,237.5 (1,282.5, 10,332.0) | 1,756.0 (779.0, 7,442.0) |
| IL-6 (pg/ml)* | 51.8 (11.3, 211.0) | 51.3 (11.8, 195.0) | 60.7 (9.5, 220.0) | 50.7 (11.7, 179.0) | 73.5 (11.3, 220.0) | 51.8 (11.9, 215.0) | 59.4 (9.5, 176.0) | 51.3 (11.8, 213.0) | 73.2 (9.5, 183.0) |
| IL-8 (pg/ml)* | 28.7 (14.2, 85.8) | 36.9 (18.2, 113.0) | 15.7 (6.6, 27.4) | 35.7 (18.1, 102.0) | 13.0 (6.6, 19.9) | 35.7 (17.6, 102.0) | 13.0 (6.6, 18.6) | 35.6 (17.5, 106.0) | 15.0 (6.6, 19.9) |
| IL-10 (pg/ml)* | 27.5 (14.0, 70.0) | 33.6 (16.9, 78.3) | 20.3 (8.0, 54.8) | 33.6 (17.9, 71.3) | 12.7 (7.1, 54.8) | 33.5 (16.5, 72.4) | 16.0 (7.1, 54.8) | 33.6 (17.1, 71.9) | 14.3 (7.1, 54.8) |
| IP-10 (pg/ml)* | 591.0 (234.0, 1,258.0) | 678.0 (256.0, 1,209.5) | 350.0 (143.0, 1,457.0) | 652.0 (258.0, 1,258.0) | 316.5 (109.0, 1,065.0) | 630.0 (248.0, 1,137.0) | 347.5 (109.0, 3,004.0) | 660.0 (256.0, 1,259.0) | 288.0 (82.4, 927.0) |
| PCT (ng/ml)* | 0.8 (0.4, 3.7) | 1.0 (0.5, 5.1) | 0.5 (0.2, 1.5) | 1.0 (0.5, 4.4) | 0.5 (0.2, 1.0) | 1.0 (0.5, 4.9) | 0.3 (0.2, 1.0) | 1.0 (0.5, 5.1) | 0.4 (0.2, 1.0) |
| sTNF-R1 (pg/ml)* | 2,121.0 (1,549.0, 3,230.0) | 2,245.5 (1,590.5, 3,790.0) | 1,752.0 (1,356.0, 2,266.0) | 2,267.0 (1,585.0, 3,603.0) | 1,746.0 (1,356.0, 1,936.0) | 2,266.0 (1,596.0, 3,548.0) | 1,580.5 (1,356.0, 1,839.0) | 2,245.0 (1,581.0, 3,575.5) | 1,740.0 (1,356.0, 1,936.0) |
| sTREM1 (pg/ml)* | 376.0 (273.0, 564.0) | 420.5 (286.0, 619.0) | 344.0 (205.0, 421.0) | 415.0 (282.0, 615.0) | 344.0 (214.0, 421.0) | 420.0 (301.0, 623.0) | 225.0 (198.0, 348.0) | 417.5 (286.0, 624.0) | 261.0 (205.0, 416.0) |
| suPAR (ng/ml)* | 5.4 (4.1, 7.7) | 5.7 (4.4, 8.0) | 4.3 (2.7, 5.6) | 5.6 (4.4, 8.0) | 3.6 (2.6, 5.8) | 5.6 (4.4, 8.1) | 3.6 (2.7, 5.0) | 5.6 (4.4, 8.1) | 3.2 (2.6, 4.8) |
| **Other laboratory markers** | | | | | | | | | |
| Lactate (mmol/l)* | 1.4 (0.8, 2.1) | 1.4 (0.9, 2.4) | 1.0 (0.6, 1.7) | 1.4 (0.9, 2.5) | 0.9 (0.6, 1.7) | 1.4 (0.9, 2.5) | 0.7 (0.6, 1.6) | 1.4 (0.9, 2.4) | 0.8 (0.6, 1.9) |
| Glucose (mmol/l)* | 6.0 (5.1, 7.1) | 6.1 (5.1, 7.3) | 5.7 (5.1, 6.8) | 6.1 (5.1, 7.3) | 5.7 (5.1, 6.8) | 6.1 (5.2, 7.3) | 5.7 (5.0, 6.8) | 6.1 (5.1, 7.2) | 5.7 (5.1, 6.8) |
| Hb (g/dL)* | 10.8 (9.5, 12.1) | 10.3 (9.3, 11.8) | 11.8 (10.9, 12.2) | 10.4 (9.3, 11.9) | 11.6 (10.6, 12.5) | 10.4 (9.3, 11.9) | 12.0 (10.9, 12.5) | 10.4 (9.3, 11.9) | 11.8 (10.6, 12.5) |
| **Prognostication** | | | | | | | | | |
| Time to event (hours)* | 6.0 (1.0, 21.0) | 3.0 (1.0, 20.5) | 9.0 (4.0, 21.0) | 4.0 (1.0, 24.0) | 9.0 (4.0, 17.0) | 4.0 (1.0, 18.0) | 10.5 (6.0, 21.0) | 4.0 (1.0, 23.5) | 10.0 (4.0, 19.0) |

[1]Median (Q1, Q3); n (%)

[a]Calculated in children aged 3-60 months (R package: *zscorer*);[33] [b]Assessed using the Alert Voice Pain Unresponsive (AVPU) scale. CRT = capillary refill time; LRTI = lower respiratory tract infection; URTI = upper respiratory tract infection.

*Missing data: wasted, n = 1; MUAC-for-age z-score, n = 41; WHO danger sign, n = 1; convulsions, n = 1; lethargy, n = 1, oxygen saturation, n = 60; ANG-1, ANG-2, sFlt-1, CRP, IL-1ra, IL-6, IL-8, IL-10, IP-10, PCT, sTNF-R1, sTREM1, n = 6; CHI3L1, lactate, glucose, n = 7; suPAR, n = 8; Hb, n = 13; time to severe disease, n = 6.

### 19. Table S16: Variation in predicted cost-effectiveness with increasing referral costs.

**Comparator: WHO danger signs**

All models were predicted to be cost-effective irrespective of referral cost (all models had better sensitivity and specificity).

**Comparator: clinical model**

The *pulse oximetry model* and the *combined model* had better specificity compared to the *clinical model.* As a result, cost-effectiveness was predicted to improve as referral costs increased. For the *sTREM1 model*, Table S7 indicates the maximum referral cost per patient at which it was predicted to remain cost-effective compared to the *clinical model*, using the two cost-effectiveness thresholds (CETs).

**Table S16: Maximum referral cost per patient at which models remain cost-effective**

| Model | Cost-effective up to maximum referral cost vs. *clinical model* (USD) | |
|---|---|---|
| | CET = $459/DALY averted | CET = $2,551/DALY averted |
| Clinical + sTREM1 | $593 | $5,217 |

**Comparator: pulse oximetry model**

The *pulse oximetry model* was predicted to be cost-effective compared to the *clinical model* with an incremental cost-effectiveness ratio (ICER) of $26/DALY averted, and due to its better specificity cost-effectiveness was predicted to improve as referral costs increased. Neither the *sTREM1* nor *combined models*, were predicted to be cost-effective compared to the *pulse oximetry model* using either of the two CETs.

### 20. Table S17. Spot Sepsis Investigator Group.

| Name | Affiliation |
|---|---|
| **Mohammad Yazid Abdad** | Mahidol-Oxford Tropical Medicine Research Unit, Mahidol University, Thailand; Centre for Tropical Medicine and Global Health, University of Oxford, UK |
| **Riris Andono Ahmad** | Centre for Tropical Medicine, Universitas Gadjah Mada, Indonesia |
| **Dinh Thi Van Anh** | Viet Nam National Children's Hospital, Viet Nam |
| **Eggi Arguni** | Centre for Tropical Medicine, Universitas Gadjah Mada, Indonesia |
| **Elizabeth A Ashley** | Lao-Oxford-Mahosot Hospital-Wellcome Trust Research Unit, Mahosot Hospital, Laos; Centre for Tropical Medicine and Global Health, University of Oxford, UK |
| **Elizabeth M Batty** | Mahidol-Oxford Tropical Medicine Research Unit, Mahidol University, Thailand; Centre for Tropical Medicine and Global Health, University of Oxford, UK |
| **Stuart D Blacksell** | Mahidol-Oxford Tropical Medicine Research Unit, Mahidol University, Thailand; Centre for Tropical Medicine and Global Health, University of Oxford, UK |
| **Latsaniphone Boutthasavong** | Lao-Oxford-Mahosot Hospital-Wellcome Trust Research Unit, Mahosot Hospital, Laos |
| **Sakib Burza** | Médecins Sans Frontières Operational Centre Barcelona, Spain; Clinical Research Department, London School of Hygiene and Tropical Medicine, UK; Health in Harmony, United States of America |
| **Arjun Chandna** | Cambodia Oxford Medical Research Unit, Angkor Hospital for Children, Cambodia; Centre for Tropical Medicine and Global Health, University of Oxford, UK; Clinical Research Department, London School of Hygiene & Tropical Medicine, UK |
| **Ngoun Chanpheaktra** | Angkor Hospital for Children, Cambodia |
| **Khalid Shams Choudhury** | Médecins Sans Frontières Operational Centre Barcelona, Spain |
| **Tran Quoc Dat** | Vietnam National Children's Hospital, Viet Nam |
| **Vu Quoc Dat** | Hanoi Medical University, Viet Nam |
| **Nicholas P J Day** | Mahidol-Oxford Tropical Medicine Research Unit, Mahidol University, Thailand; Centre for Tropical Medicine and Global Health, University of Oxford, UK |
| **Arjen M Dondorp** | Mahidol-Oxford Tropical Medicine Research Unit, Mahidol University, Thailand; Centre for Tropical Medicine and Global Health, University of Oxford, UK |
| **Prakash Ghosh** | International Centre for Diarrhoeal Disease Research, Bangladesh; Institute of Animal Hygiene and Veterinary Public Health, Leipzig University, Germany; Technische Universität Berlin, Germany |
| **Carolina Jimenez** | Médecins Sans Frontières Operational Centre Barcelona, Spain |
| **Kevin Kain** | Department of Laboratory Medicine and Pathobiology, University of Toronto, Canada |
| **Muhammad Karyana** | INA-RESPOND, Ministry of Health Republic Indonesia, Indonesia |
| **Suy Keang** | Angkor Hospital for Children, Cambodia; Cambodia Oxford Medical Research Unit, Angkor Hospital for Children, Cambodia |
| **Sommay Keomany** | Salavan Provincial Hospital, Laos |
| **Rungnapa Khamboocha** | Mahidol-Oxford Tropical Medicine Research Unit, Mahidol University, Thailand |

| | |
|---|---|
| **Constantinos Koshiaris** | Department of Primary Care Health Sciences, University of Oxford, UK |
| **Khamfong Kunlaya** | Lao-Oxford-Mahosot Hospital-Wellcome Trust Research Unit, Mahosot Hospital, Laos |
| **Estrella Lasry** | Médecins Sans Frontières Operational Centre Barcelona, Spain |
| **Bui Thanh Liem** | University of Medicine and Pharmacy at Ho Chi Minh City, Vietnam |
| **Nguyen Huy Luan** | University of Medicine and Pharmacy at Ho Chi Minh City, Vietnam |
| **Yoel Lubell** | Mahidol-Oxford Tropical Medicine Research Unit, Mahidol University, Thailand; Centre for Tropical Medicine and Global Health, University of Oxford, UK; Amsterdam Institute of Global Health and Development, The Netherlands |
| **Raman Mahajan** | Médecins Sans Frontières Operational Centre Barcelona, Spain |
| **Saysamone Malavong** | Savannakhet Provincial Hospital, Laos |
| **Mayfong Mayxay** | Lao-Oxford-Mahosot Hospital-Wellcome Trust Research Unit, Mahosot Hospital, Laos; Institute for Research and Education Development, University of Health Sciences, Laos; Centre for Tropical Medicine and Global Health, University of Oxford, UK |
| **Chonticha Menggred** | Mahidol-Oxford Tropical Medicine Research Unit, Mahidol University, Thailand |
| **Dinesh Mondal** | International Centre for Diarrhoeal Disease Research, Bangladesh |
| **Phung Nguyen The Nguyen** | University of Medicine and Pharmacy at Ho Chi Minh City, Vietnam |
| **Chris Painter** | Lao-Oxford-Mahosot Hospital-Wellcome Trust Research Unit, Mahosot Hospital, Laos; Mahidol-Oxford Tropical Medicine Research Unit, Mahidol University, Thailand; Centre for Tropical Medicine and Global Health, University of Oxford, UK |
| **Rafael Perera-Salazar** | Department of Primary Care Health Sciences, University of Oxford, UK |
| **Chom Phaiphichit** | Lao-Oxford-Mahosot Hospital-Wellcome Trust Research Unit, Mahosot Hospital, Laos |
| **Chanthala Phamisith** | Savannakhet Provincial Hospital, Laos |
| **Phan Huu Phuc** | Viet Nam National Children's Hospital, Viet Nam |
| **Tiengkham Pongvongsa** | Savannakhet Provincial Health Office, Laos |
| **Sayaphet Rattanavong** | Lao-Oxford-Mahosot Hospital-Wellcome Trust Research Unit, Mahosot Hospital, Laos |
| **Michael Rekart** | Médecins Sans Frontières Operational Centre Barcelona, Spain |
| **Melissa Richard-Greenblatt** | Centre for Tropical Medicine and Global Health, University of Oxford, UK; Department of Laboratory Medicine and Pathobiology, University of Toronto, Canada; Department of Pediatric Laboratory Medicine, The Hospital for Sick Children, Canada |
| **Bran Sambou** | Cambodia Oxford Medical Research Unit, Angkor Hospital for Children, Cambodia |
| **Mohammad Shomik** | International Centre for Diarrhoeal Disease Research, Bangladesh |
| **Phouthalavanh Souvannasing** | Salavan Provincial Hospital, Laos |
| **Phattaranit Tanunchai** | Mahidol-Oxford Tropical Medicine Research Unit, Mahidol University, Thailand |
| **Janjira Thaipadungpanit** | Mahidol-Oxford Tropical Medicine Research Unit, Mahidol University, Thailand; Faculty of Tropical Medicine, Mahidol University, Thailand |

| | |
|---|---|
| **Watcharintorn Thongpiam** | Mahidol-Oxford Tropical Medicine Research Unit, Mahidol University, Thailand |
| **Bang Huyen Tran** | Oxford University Clinical Research Unit, Ho Chi Minh City, Viet Nam |
| **Claudia Turner** | Angkor Hospital for Children, Cambodia; Cambodia Oxford Medical Research Unit, Angkor Hospital for Children, Cambodia; Centre for Tropical Medicine and Global Health, University of Oxford, UK |
| **Paul Turner** | Cambodia Oxford Medical Research Unit, Angkor Hospital for Children, Cambodia; Centre for Tropical Medicine and Global Health, University of Oxford, UK |
| **Souphaphone Vannachone** | Lao-Oxford-Mahosot Hospital-Wellcome Trust Research Unit, Mahosot Hospital, Laos |
| **Asama Vinitsorn** | Mahidol-Oxford Tropical Medicine Research Unit, Mahidol University, Thailand |
| **Ranitha Vongpromek** | Mahidol-Oxford Tropical Medicine Research Unit, Mahidol University, Thailand |
| **Manivanh Vongsouvath** | Lao-Oxford-Mahosot Hospital-Wellcome Trust Research Unit, Mahosot Hospital, Laos |
| **Naomi Waithira** | Mahidol-Oxford Tropical Medicine Research Unit, Mahidol University, Thailand; Centre for Tropical Medicine and Global Health, University of Oxford, UK |
| **James A Watson** | Mahidol-Oxford Tropical Medicine Research Unit, Mahidol University, Thailand; Centre for Tropical Medicine and Global Health, University of Oxford, UK |
| **Mikhael Yosia** | Médecins Sans Frontières Operational Centre Barcelona, Spain |
| **Asri Yuniastuti** | Wates District Hospital, Indonesia |
